# Adult vaccination in Brazil: a cross-sectional survey on physicians’ prescription habits

**DOI:** 10.1101/2021.02.02.21251016

**Authors:** Fernando B Serra, Diogo Ribeiro, Paula M Batista, Thais N F Moreira

**Author notes:** **Corresponding author: Thais das Neves Fraga Moreira**, MSD Brazil, Avenida Chucri Zaidan, 296, Vila Cordeiro – São Paulo/SP – Brazil - ZIP CODE : 04583-110. **Authors’ emails and phone numbers**: Fernando B Serra –; (55)11 5189 7584 Diogo Ribeiro –; (351) 21 382 54 40 Paula M Batista –; (55) 11 5189 7679 Thais N F Moreira –; (55) 11 5189 7204.

## Abstract

**OBJECTIVE:** To characterize adult and older adult vaccination practices of physicians, from various medical specialties, in Brazil; identify barriers influencing prescription of vaccines to these populations, and evaluate the physicians’ knowledge on routinely prescribed vaccines.

**METHODS:** Cross-sectional survey conducted in Brazil between June-August 2018. Eligible physicians included those from general practice/family medicine, geriatrics, cardiology, gynecology, endocrinology, infectious disease and pulmonology. The survey’s questions addressed the physicians’ prescription habits, sociodemographic and clinical practice characteristics, barriers to vaccines’ prescription, and physicians’ knowledge regarding routinely prescribed vaccines. The study focused on the vaccines recommended by the Brazilian Society of Immunization (SBIm) for adults and older adults (years 2017-2018). Study sample was stratified according to the number of physicians per specialty and Brazilian region.

**RESULTS:** A total of 1068 surveys were completed. The vaccines prescribed by the highest proportions of physicians were Influenza (>90% of physicians for adults and older adults), Hepatitis B (adults: 87%; older adults: 59%) and Yellow Fever (adults: 77.7%; older adults: 58.5%). Underprescription was reported by less than 20% of prescribing physicians for all adult and older adult recommended vaccines. The most common barriers to vaccination were the high vaccine cost, lack of time during appointments and lack of patient interest. Knowledge on target populations, dosage schedule and availability in the Unified Public Health System (SUS) was generally low.

**CONCLUSIONS:** The results showed a considerable variability of prescribing habits across recommended vaccines. Although most prescribing physicians seem to be aware of the importance of adult and older adult vaccination, knowledge deficits on vaccines’ target populations, dosage schedule and availability in the SUS may hamper their ability to prescribe vaccines to all patients with an indication.

## INTRODUCTION

Vaccines are recommended throughout a person’s lifetime to avert vaccine-preventable diseases (VPDs) and their complications^1^. In Brazil, the National Immunization Program (NIP) supplies free-of-charge vaccines to all age groups as part of a defined immunization schedule^2^. Currently, five vaccines are provided to adults (Influenza, Hepatitis B, Yellow Fever, Tetanus and diphtheria [Td], and Measle, mumps and rubella [MMR]), of which four are also provided to older adults (all except MMR)^3^. In addition, the Brazilian Society of Immunization (SBIm) annually issues immunization schedules for adults (20-59 years)^4^, older adults (≥60 years)^5^, and special populations at increased risk of VPDs^6^.

Age-related disorders and comorbid conditions, such as diabetes, cardiovascular diseases, and cancer increase the risk and severity of VPDs^7–9^. Acute infectious diseases can also reduce functional capacity in older adults in the short and long term^10^. Thus, attaining an appropriate vaccination coverage represents an important public health strategy for healthy aging, capable of reducing mortality, morbidity and of improving quality of life among these patients^7, 9, 11^.

Studies have reported that adult vaccination coverage rate (VCR) is still at a suboptimal level in various countries^12, 13^, including Brazil^14, 15^. This has been attributed to limited public awareness, misinformation, vaccine hesitancy, gaps in recommending vaccines during healthcare visits and vaccine costs^1, 16^. In Brazil, the target for Influenza VCR (80%), which is delivered in mass campaigns supported by strong media communication, has been consistently met since 2011 in older adults^17^. However, a considerably lower coverage has been reported among patients with diabetes (47.2% in 2015)^14^. Furthermore, studies have also shown inadequate pneumococcal VCR in adults and older adults (50.8%)^15^, especially in diabetic patients (26.1%)^14^.

Detailed understanding of factors responsible for low VCR among adult populations is required to establish effective immunization strategies. Studies on physicians’ behavior toward adult vaccination are presently lacking, despite the fact that physicians’ recommendations represent the leading factor to influence patients’ adherence ^18, 19^. Thus, this study aimed to characterize vaccination practices of physicians from various medical specialties in Brazil (general practice (GP)/family medicine (FM), geriatrics, cardiology, gynecology, endocrinology, infectious disease and pulmonology). We also intended to identify the barriers influencing vaccine prescriptions and physicians’ knowledge regarding routinely prescribed vaccines.

## METHODS

This was a national, cross-sectional study conducted between June and August 2018 in Brazil. Study data were collected through an online survey self-completed by eligible physicians. The survey’s questions addressed the physicians’ prescription habits, sociodemographic and clinical practice characteristics, barriers to the prescription of vaccines to adult and older adult patients, and physicians’ knowledge regarding routinely prescribed vaccines. This study focused exclusively on the vaccines recommended by the SBIm for adults and older adults in the years 2017/2018^4, 5^ – Box 1.

**Box 1.**
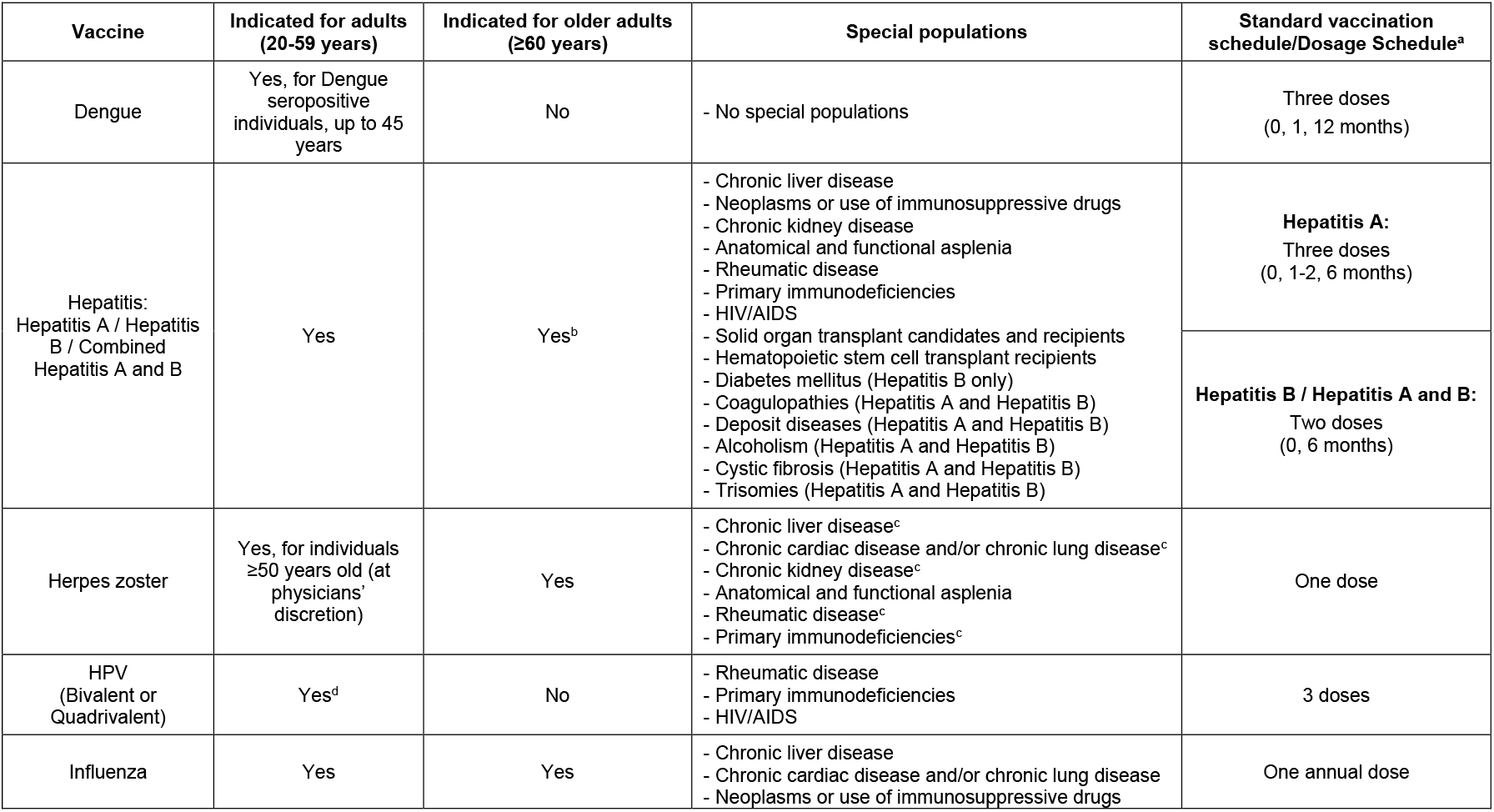
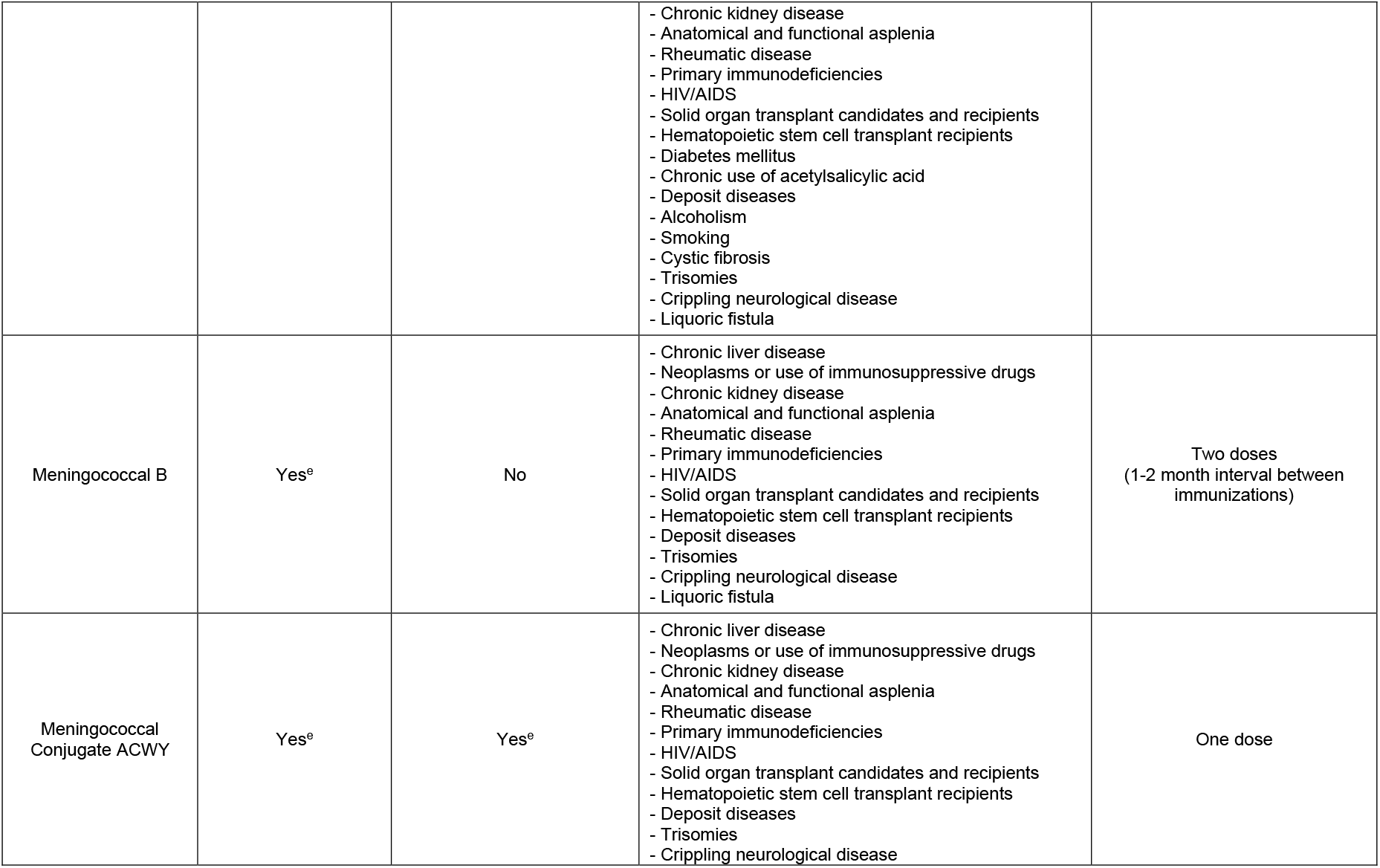
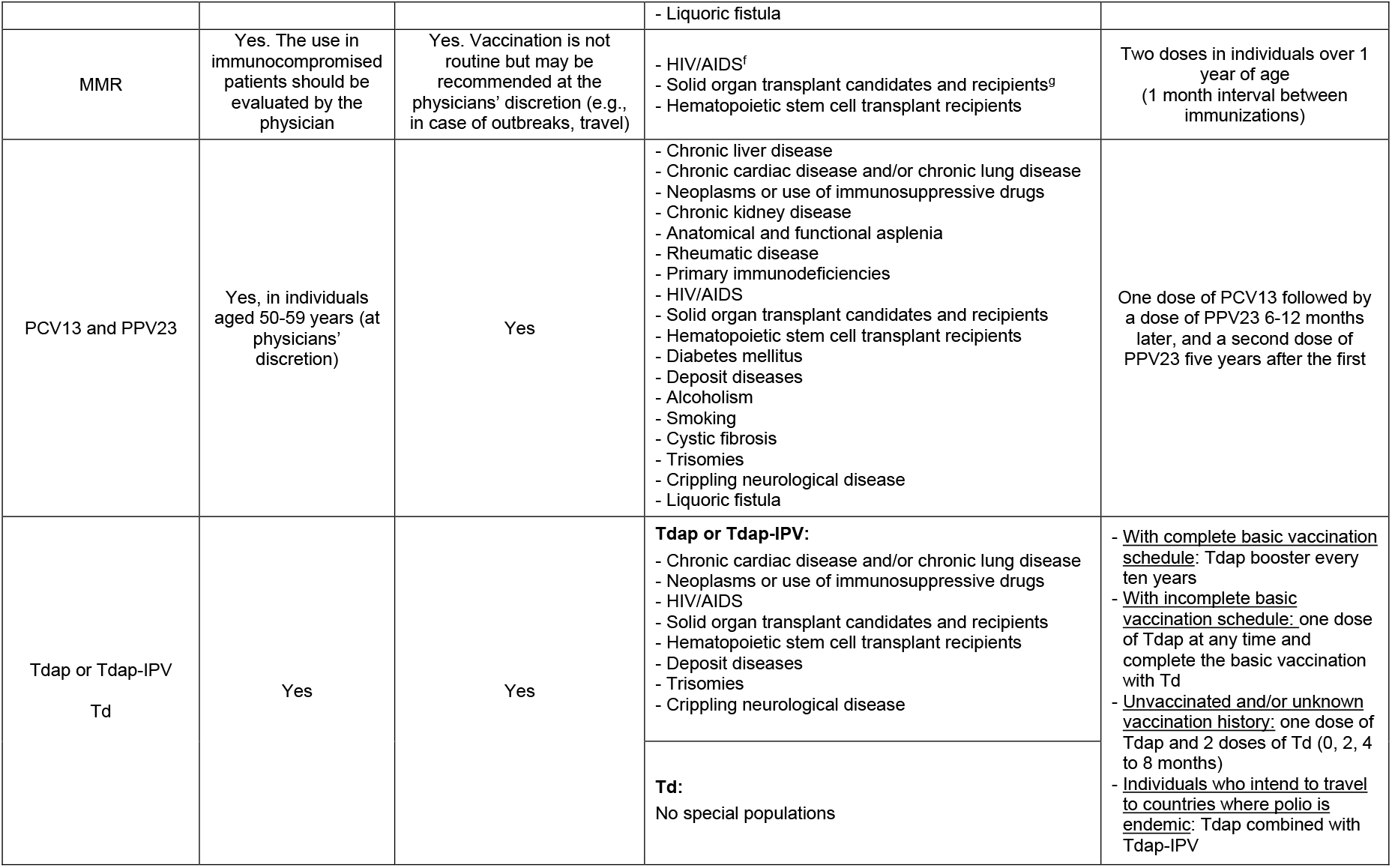
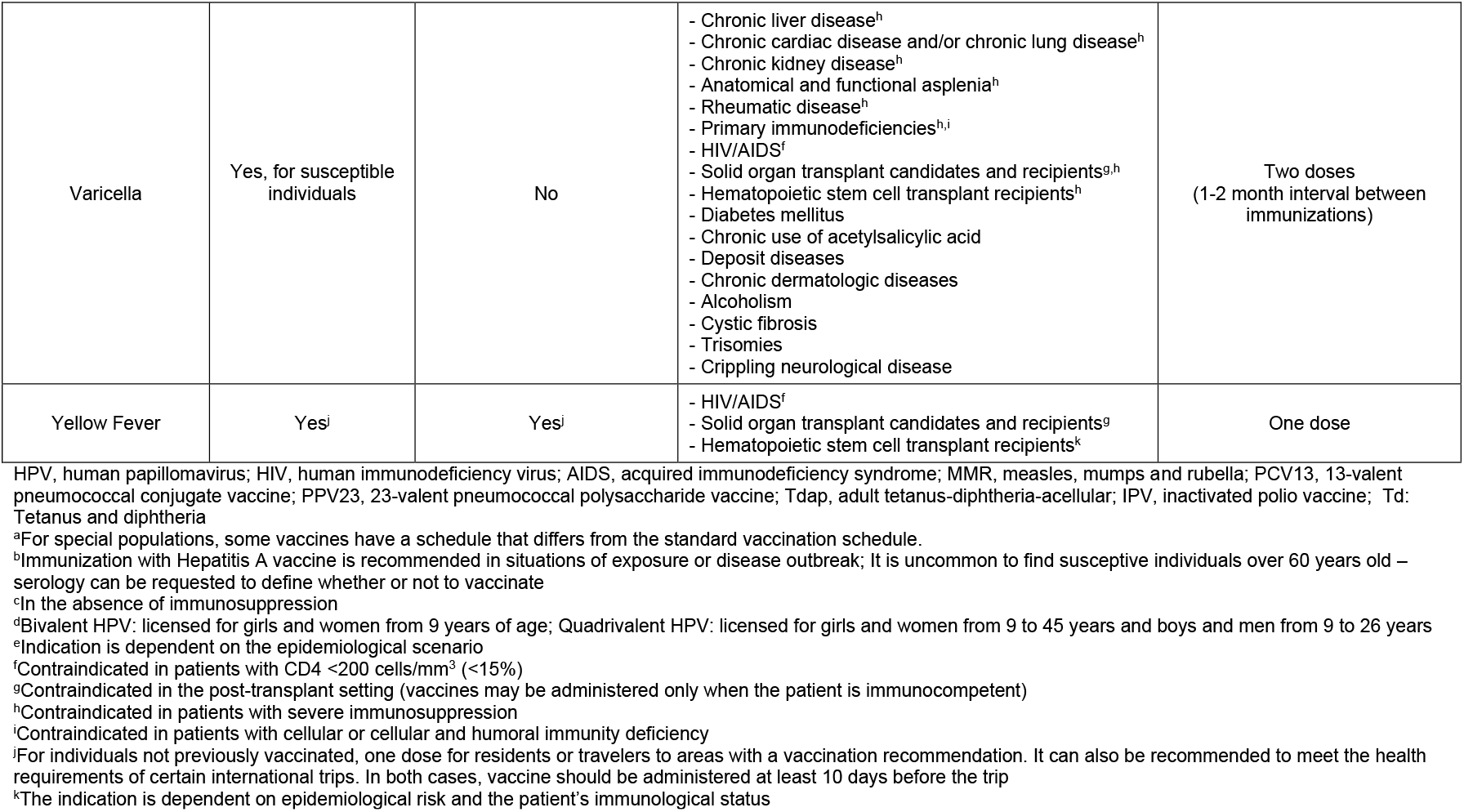
Brazilian Society of Immunization’s recommendations for adults and older adults at the time of the study (2017-2018)^4, 5, 20^

This study was approved by the Ethics Committee of Universidade Federal de São Paulo.

### Physicians’ prescription habits

In terms of prescription habits, the survey collected the information depicted in Figure 1 for each adult and older adult recommended vaccine.

**Figure 1.**
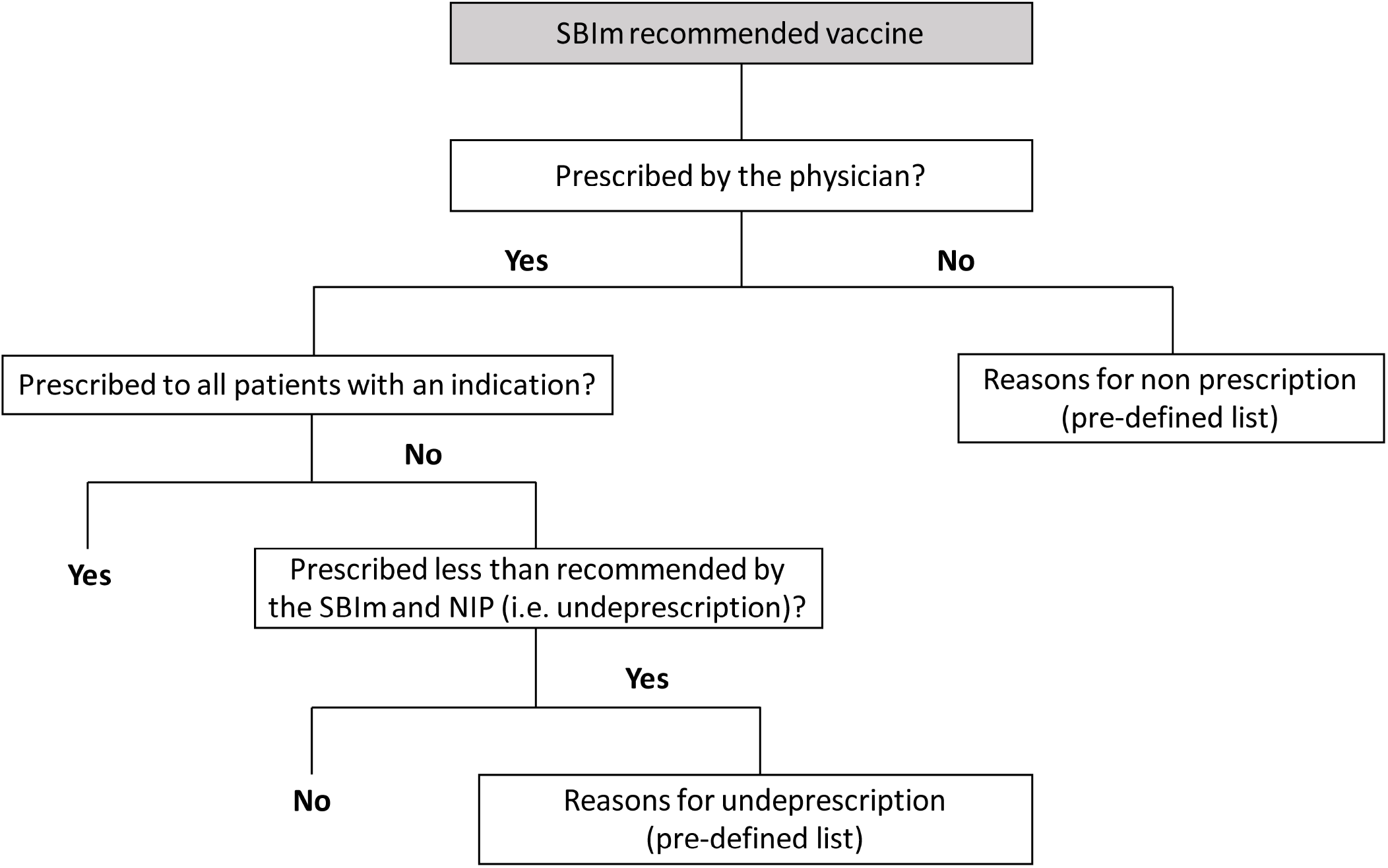
Information collected on prescription habits for adult and older adult recommended vaccines. SBIm, Brazilian Society of Immunization; NIP, National Immunization Program

Figure 1 – Information collected on prescription habits for adult and older adult SBIm recommended vaccines

Underprescription was defined as prescribing less than recommended by the SBIm and NIP. The survey did not provide the immunization recommendations from these entities and responding physicians were expected to use their own knowledge of these recommendations to identify and report underprescription.

### Physicians’ knowledge on routinely prescribed vaccines

The survey assessed the physicians’ knowledge regarding the recommended vaccines’ target populations, dosage schedule, and availability in SUS. We used the most up-to-date immunization schedules issued by the SBIm for adults^4^ and older adults^5^ (2017-2018) and special populations (2015-2016)^20^ at the time of the study to define the correct answers. For availability in SUS, the correct answers for 23-valent pneumococcal polysaccharide vaccine (PPV23) and quadrivalent human papillomavirus (HPV) vaccine considered more recent information published by the Ministry of Health instead^21, 22^. All correct answers are detailed in Supplementary Table 1 (for target populations), Table 2 (for dosage schedule) and Table 3 (for availability in SUS).

For target populations, physicians should select the age groups (Adults [20-59 years] and/or Older adults [>60 years]) for which each vaccine is recommended, as well as the respective special populations. The physicians’ knowledge on this topic was assessed by means of an overall score. This score, ranging from zero to one, was calculated based on the individual scores obtained by each physician in the question regarding target populations. The individual score corresponded to the ratio between the number of correct options selected by the physician and the total number of correct answers defined for each recommended vaccine. Each incorrect option selected nullified a correct one. The only exception to this rule occurred when a SBIm recommendation did not encompass all patients within a given subpopulation (e.g., varicella vaccine is recommended for patients with HIV/AIDS, but not for those with CD4 <200 cells/mm^3^). In these cases, the option pertaining to the special population was considered correct, but did not nullify a correct one if not selected.

Physicians’ knowledge on dosage schedules and availability in the SUS was assessed by determining the proportions of correct answers in the respective survey questions. The correct answers for availability in the SUS focused on whether the vaccines were available free of charge at the basic health units (BHUs).

### Sample size

A sample of 1068 physicians was needed to determine the proportion of physicians prescribing each SBIm recommended vaccine, with a margin of error of 3% and a significance level of 0.05. In the absence of data on the proportion of prescribing physicians for SBIm recommended vaccines, we followed the most conservative approach by considering a proportion of 50%.

The sample was stratified according to the number of active physicians per Brazilian region and medical specialty (Supplementary Table 4). This was based on the number of active physicians in Brazil (overall and per region) according to the Federal Council of Medicine’s website^23^ and on the distribution of physicians across medical specialties of interest, as described by Scheffer M. et al^24^ – see Supplementary Material for a detailed description of this process.

### Recruitment of physicians

The recruitment of physicians was based on the Fine Panel, a large physicians’ panel covering Brazil and other Latin America countries. The recruitment of physicians for this is based on various sources: public information and websites, contacts of affiliated physicians provided by hospitals and medical associations, contacts obtained during visits to hospitals and clinics, voluntary registrations and peer-to-peer recommendations from registered physicians. Physicians were eligible to complete the survey if they had a valid registration in the Fine Panel, either before or after being invited to participate in the study. Additionally, physicians had to meet all the following criteria: 1) worked at a public and/or private medical institution in Brazil; 2) belonged to one of the medical specialties of interest; 3) saw at least 30 patients per week. Conversely, physicians were not eligible if a sufficient number of physicians from the same medical specialty and Brazilian region had already completed the survey.

### Statistical Analysis

Qualitative variables were summarized by absolute and relative frequencies. For the proportion of physicians prescribing each recommended vaccine, 95% confidence intervals were also computed. Quantitative variables were summarized by descriptive statistics, namely mean, median, standard deviation, minimum and maximum.

## RESULTS

### Study sample construction

A total of 76 104 physicians were invited to participate in the study to obtain 1068 completed surveys. Of these, 759 surveys were completed by physicians registered in the Fine Panel before being invited to participate (8 741 invitations), while the remaining 309 were completed by physicians who registered after receiving the invitation (67 364 invitations). The characteristics of the general population of physicians in Brazil, those invited to participate in the study, and those completing the survey are described in Supplementary Table 13.

### Sociodemographic and clinical practice characterization

Almost half of the physicians (48.2%) were gynecologists – Table 1. On average, physicians had been practicing their medical specialty for 17.1 (±10.0) years and prescribing vaccines to adults and/or older adults for 13.9 (±8.3) years. Most physicians (89.9%) routinely recommended vaccines.

**Table 1.** – Participants’ sociodemographic and clinical practice characteristics

|  | **Total (n=1068)** |
| --- | --- |
| Age (years) | 44.70 ± 9.91 |
| Sex (female) | 574 (53.7) |
| Brazilian region |  |
| North | 48 (4.5) |
| Northeast | 191 (17.9) |
| Midwest | 88 (8.2) |
| Southeast | 580 (54.3) |
| South | 161 (15.1) |
| Medical specialty best defining clinical practice |  |
| Cardiology | 244 (22.8) |
| General Practice/Family Practice | 85 (8.0) |
| Endocrinology | 80 (7.5) |
| Geriatrics | 26 (2.4) |
| Gynecology | 515 (48.2) |
| Infectious Disease | 59 (5.5) |
| Pulmonology | 59 (5.5) |
| Place(s) of work |  |
| Public hospital | 569 (53.3) |
| Private hospital | 604 (56.5) |
| Public medical office | 367 (34.4) |
| Private medical office for patients with health insurance plans | 838 (78.5) |
| Private medical office (payment is made exclusively by the patient) | 620 (58.0) |
| Years of medical specialty practice<sup>a</sup> (years) (n=1063) | 17.06 ± 9.96 |
| Number of patients cared for during a regular work week | 113.09 ± 76.72 |
| Routinely recommends vaccines to patients | 959 (89.8) |
| Vaccine prescription as part of medical practice (n=959) | 944 (98.4) |
| Characterization of vaccine prescription in clinical practice (n=944) |  |
| Very common | 383 (40.6) |
| Common | 457 (48.4) |
| Not very common | 104 (11.0) |
| Time prescribing vaccines to adult and older adult patients (years) (n=955) | 13.85 ± 8.28 |
| Vaccines prescribed per week to adult and older adult patients |  |
| None | 62 (5.8) |
| 1-5 | 388 (36.3) |
| 6-10 | 279 (26.1) |
| 11-20 | 220 (20.6) |
| 21-50 | 92 (8.6) |
| >50 | 27 (2.5) |

Results are displayed as n (%) or mean ± standard deviation.
N=1068 unless otherwise specified.
All summary statistics include only non-missing values.
<sup>a</sup>Excluding years of medical residency/specialization.

### Prescription habits

The proportions of physicians prescribing each recommended vaccine (to all or some of their patients with an indication) are summarized in Table 2.

**Table 2.**
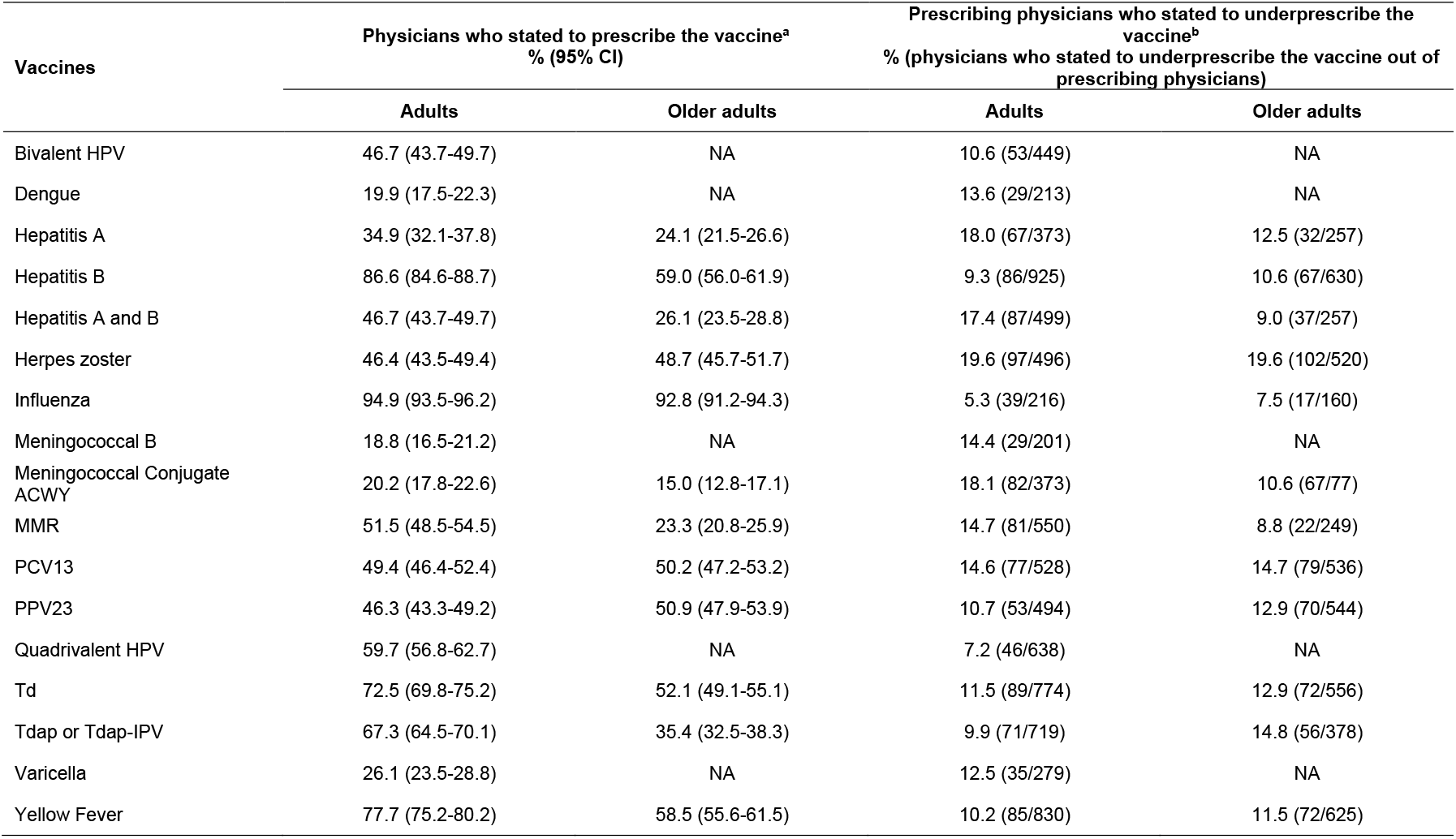
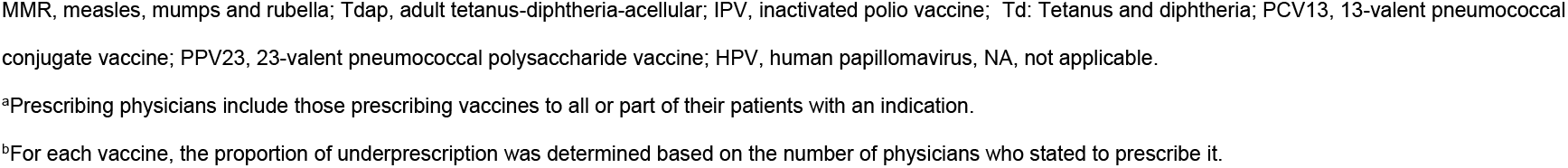
– Physicians prescribing and underprescribing vaccines recommended by the Brazilian Society of Immunization to adult and older adults

The vaccines prescribed by the highest proportions of physicians to adults and older adults were Influenza (≥90% of physicians in both groups), Hepatitis B (87% for adults and 59% for older adults) and Yellow Fever (77.7% for adults and 58.5% for older adults). Conversely, Meningococcal B (18.8%) and Meningococcal conjugated ACWY (20.2%) were prescribed by the lowest proportions of physicians for adults and older adults, respectively.

### Prescription of vaccines to all patients with an indication

The proportion of prescribing physicians who stated to prescribe vaccines to all adult and older adult patients with an indication were above 75% for all recommended vaccines – Supplementary Table 5. Influenza was the vaccine prescribed by the highest proportion of physicians to all adult (93.3%) and older adult (91.6%) patients with an indication. Zoster vaccine had the lowest proportions of physicians prescribing to all patients with an indication (76% and 76.9% for adults and older adults, respectively).

### Barriers to vaccine prescription

#### Reasons for non-prescription

The most frequently selected reason for non-prescription was the fact that the vaccine did not belong to the scope of the physicians’ medical specialty. Doubts regarding the vaccine’s indication and fear of side effects were the second most frequently selected reasons for non-prescription for nine and eight of the 17 adult recommended vaccines, respectively. For each vaccine, the reasons leading to non-prescription in adults and older adults are provided in Supplementary Tables 6 and 7, respectively.

### Reasons for underprescription

The proportions of prescribing physicians who stated to underprescribe vaccines were below 20% for all adult and older adult recommended vaccines – Table 2.

The high vaccine cost and lack of time during the appointment were the barriers most frequently selected by physicians for adult recommended vaccines (6 and 5 vaccines, respectively). For older adult recommended vaccines, the most commonly selected barriers were the high vaccine cost and lack of patient interest (4 vaccines each).

For each vaccine, the reasons leading to underprescription in adults and older adults are presented in Supplementary Tables 8 and 9, respectively.

### Physicians’ knowledge on recommended vaccines

Figure 2 shows the result on physicians’ knowledge regarding all recommended vaccines per target populations. Td and Meningococcal B were the vaccines for which physicians presented the highest and lowest knowledge, respectively (mean score of 0.76 and 0.09, respectively).

**Figure 2.**
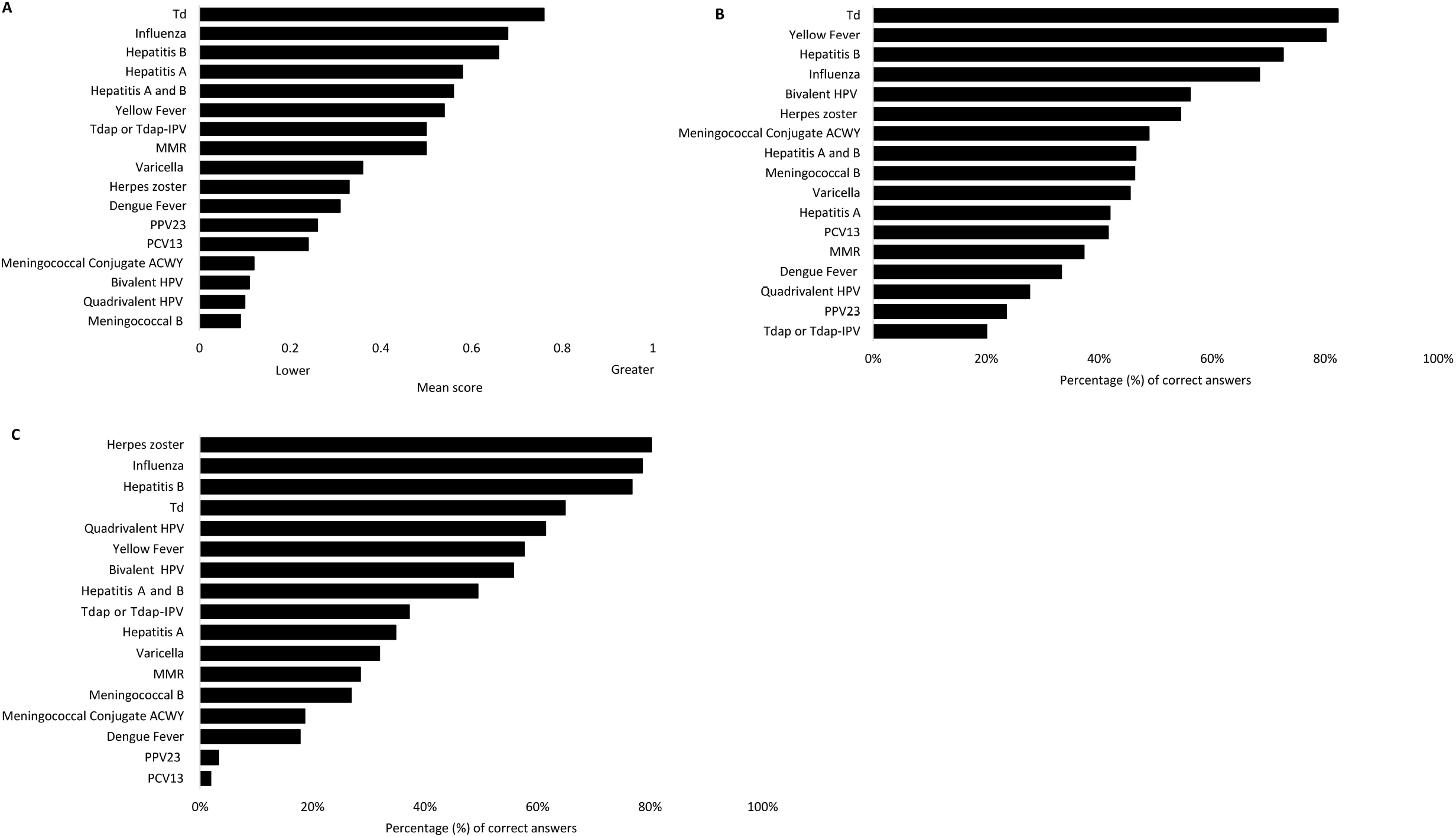
Knowledge on SBIm recommended vaccines’ target populations (A), dosage schedule (B) and availability in the SUS (C). Td, Tetanus and diphtheria; Tdap, adult tetanus-diphteria-accelullar; IPV, inactivated polio vaccine; MMR, Measles, Mumps, and Rubella; PPV23, 23-valent pneumococcal polysaccharide vaccine; PCV13, 13-valent pneumococcal conjugate vaccine; HPV, human papillomavirus

Regarding dosage schedule, Zoster was associated with the highest knowledge (80.2% of correct answers). Conversely, the lowest percentage of correct answers on this question was observed for PCV13 (1.9%).

Concerning availability in Brazil’s SUS, Td and adult tetanus-diphtheria-acellular (Tdap) or Tdap-inactivated polio vaccine (IPV) were the vaccines for which physicians obtained the highest and lowest percentages of correct answers, respectively (82.3% and 20.1% of correct answers, respectively).

### Knowledge by medical specialty

Results on vaccine knowledge regarding target populations, dosage schedule, and availability in the SUS by medical specialty are presented in Supplementary Tables 10, 11 and 12, respectively. The infectious disease specialists showed higher mean scores of knowledge on target populations for most vaccines, except for Hepatitis A, Tdap or Tdap-IPV, PVC13 (higher mean scores in pulmonologists) and Td (higher mean score in endocrinologists). When compared with other specialties, the proportion of correct answers regarding dosage schedule and availability in the SUS were descriptively higher among infectious disease specialists for ten and seven, respectively, of the 17 SBIm recommended vaccines.

## DISCUSSION

The burden of VPDs is increasing among older individuals with the population aging^25, 26^. Vaccination is a key and effective public health strategy to reduce adult morbidity and mortality^7, 25, 27^. However, studies conducted in Brazil show low VCR for some VPDs (e.g., pneumococcal disease)^15^, particularly among patients with comorbidities^14^. As physicians play a critical role in promoting vaccine uptake^18, 19^, characterizing their behavior towards adult and older adult vaccination is both timely and important. To the best of our knowledge, this was the first study exploring the physicians’ prescription habits and knowledge regarding adult and older adult vaccination in Brazil.

The proportions of prescribing physicians varied greatly across SBIm recommended vaccines. The low proportions of physicians prescribing the meningococcal and dengue vaccines are in line with the SBIm recommendations and may be explained, at least in part, by the fact that they are not routinely recommended for all adults. Indeed, the SBIm states that the indication for the meningococcal vaccines is dependent on the epidemiological scenario, while the dengue vaccine is recommended only for dengue seropositive individuals^4, 5^. The high proportion of prescribing physicians for Yellow Fever vaccine is likely a result of the survey conducted during the Yellow Fever epidemic in Brazil (2016-2019)^28^. It’s also worth noting that the five vaccines recommended by NIP (Influenza, Hepatitis B, Yellow Fever, MMR, and Td) were also prescribed by the highest proportions of physicians. This may suggest that the official recommendations have a greater influence on vaccine prescription than those issued by SBIm.

Though the risk groups for Influenza and pneumococcal disease are similar, the results show that a markedly higher proportion of physicians prescribe the former, both to adults and older adults. A nationwide cohort study of the non-institutionalized Brazilian population estimated that appreciable proportions of adults aged ≥50 years present risk factors for Influenza and pneumococcal disease. Indeed, approximately 16% of adults in this age group are diabetic, 12% have heart problems, 5% have cancer and 4.5% suffer from chronic kidney disease^29^. Thus, reducing the discrepancy between the proportions of physicians prescribing the Influenza and pneumococcal vaccines, by considerably increasing the latter, could prove beneficial to the life of millions of Brazilians. The importance of increasing the coverage of pneumococcal vaccines in the adult population has been highlighted in a study conducted at a tertiary care in Porto Alegre in patients aged 33-72 years. This study showed that approximately half (50.8%) of invasive pneumococcal disease cases that required hospital admission could have potentially been prevented through vaccination with PCV13 plus PPV23^15^.

The study results suggest that prescribing physicians are aware of the importance of adult and older adult vaccination. This is reflected by the fact that more than 3/4 of prescribing physicians stated to prescribe all recommended vaccines to every patient with an indication. Still, our findings also show that knowledge on vaccines’ indication is not well disseminated among interviewed physicians and that different physicians may have a distinct understanding of the same vaccine’s indication – Supplementary Table 10. Consequentially, physicians who did not accurately know a given vaccine’s indication may have incorrectly stated to prescribe it to all patients with an indication. Moreover, doubts regarding vaccines’ indication were an important factor leading to non-prescription. As such, though physicians seem mindful of the value of adult and older adult vaccination, lack of knowledge on vaccines’ indication may lead to missed opportunities to vaccinate potential vaccine recipients and increase vaccination coverage. Initiatives aimed at boosting physicians’ knowledge on recommended vaccines’ indications could thus prove valuable to increase the prescription of adult and older adult recommended vaccines in Brazil.

Though the present study focused on physicians’ habits and perceived barriers to vaccination, it’s interesting to note that lack of patient interest was among the main physician-reported barriers leading to underprescription in adults and older adults. This is in line with previous studies that explored the reasons for lack of vaccine uptake among these populations in Brazil^30, 31^. Moreover, this finding indicates that vaccine hesitancy – the delay in vaccination schedule or refusal to receive recommended vaccines, despite their availability in health services – should be a cause for concern in Brazil^31^.

Knowledge on target populations, dosage schedule and availability in the SUS was generally low. As nearly all correct answers were defined based on the SBIm immunization schedules, one possible explanation for these results is that physicians do not routinely follow the SBIm recommendations.

The results also suggest that there may be a difficulty in incorporating newer vaccines (i.e., more recently approved) into routine clinical practice. Indeed, the study data show that physicians are more familiarized with older vaccines, such as Influenza (approved in 1997)^32^, Hepatitis B (approved in 1993)^33^, and Td (used since 1975)^34^. This is reflected by the fact that these were among the four vaccines prescribed by the highest proportions of physicians to both adults and older adults and for which physicians presented the highest knowledge. Conversely, the most recently approved vaccines in Brazil (Dengue and Meningococcal B, approved in 2015^35, 36^, and Meningococcal Conjugate ACWY, approved in 2014^37^) were prescribed by the lowest proportions of physicians and were generally among those associated with the lowest knowledge, particularly for target populations and dosage schedule. In addition to not being routinely recommended for all adults, inadequate knowledge on these vaccines may have also contributed to the low proportions of prescribing physicians observed. This is supported by the fact that doubts regarding indication was the second most commonly selected reason leading to non-prescription of more recently approved vaccines (Supplementary Table 6). Thus, these results highlight the need for initiatives aimed at promoting physicians’ knowledge on SBIm recommended vaccines to focus on the most recently approved ones.

A recent study evaluated the overall coverage of NIP provided vaccines between 1994 and 2019 and explored differences in coverage between Brazilian regions^38^. The Midwest region showed the highest coverage rate (77.1%), approximately 3.6% above the national average of 73.5%. All remaining regions showed similar coverages, ranging from 72.9% (Southeast) to 73.6% (North). Though evaluating different vaccination practices across Brazilian regions was not an objective of this study, it’s worth noting that more than half (54%) of participating physicians were from the Southeast region (reflecting health care professionals geographic concentration, in line with the planned stratification), while only 8% were from the Midwest.

This study has several limitations. On one hand, the generalizability of our findings is limited by three main factors. First, only seven medical specialties were included in the study. Second, although responding physicians were stratified by medical specialty and Brazilian region (Supplementary Table 13), the stratification of the study sample did not encompass other relevant characteristics, which may have differed between responding physicians and the general population of physicians in Brazil. For instance, the study population was comprised of a considerably lower proportion of physicians working exclusively in the public sector (4.8% vs. 21.6% in the Brazilian population of physicians)^24^. Third, the study sample may have been comprised of physicians particularly interested in the topic of adult and older adult vaccination, as suggested by the high number of invitations (76 104) required to obtain 1068 surveys and by the fact that most physicians (89.8%) stated to routinely recommend vaccines. On the other hand, limitations related to the survey design and the methodology followed to define correct answers may have impacted the results obtained and should be considered during their interpretation. First, underprescription was defined as prescribing less than recommended according to the SBIm and NIP. However, the SBIm and NIP differ in the number of vaccines recommended to adults (5 versus 17, respectively) and older adults (4 versus 12, respectively). Thus, a physician may be compliant with the NIP recommendations while simultaneously prescribing less than recommended by the SBIm. As a result, underprescription may have been underestimated – for instance, physicians prescribing a given vaccine less than recommended by the SBIm may have stated not to underprescribe it if they considered the NIP recommendations only. Additionally, no immunization recommendations from SBIm and the NIP were provided in the survey. As physicians were expected to use their own knowledge on these recommendations to identify and report underprescription, those not aware of their full extent may have failed to do so. Second, physicians’ knowledge on target populations may have been overestimated for vaccines recommended for both adults and older adults (except for PCV13 and PPV23). This is because physicians who selected “All patients (regardless of age and risk group)” as the target population were given the total score of one point, even without specifying any special population according to the SBIm immunization schedule^20^ – Supplementary Table 1. Third, the correct answers regarding availability in the SUS focused on whether the vaccines were available free of charge at the BHU according to the SBIm immunization schedules^4, 5^. Nonetheless, certain vaccines are available free of charge at the Special Immunobiological Reference Centers (CRIES), which are a part of the SUS. Physicians may have thus considered the availability of vaccines in the CRIES when responding to this question, which may have impacted the results.

In conclusion, a considerable variability of prescribing habits was observed in this study across SBIm recommended vaccines. Although most prescribing physicians seem to be aware of the importance of adult and older adult vaccination, knowledge deficits on vaccines’ target populations, dosage schedule, and availability in the SUS may hamper their ability to prescribe vaccines to all patients with an indication. Initiatives to increase the physicians’ knowledge (particularly on recently approved vaccines) are recommended to increase the VCR among the adult and older adult Brazilian populations, as is the adoption of measures to tackle the most common barriers to vaccination (high cost, lack of time during appointments, and lack of patient interest).

## Data Availability

not applicable.

## Acknowledgements

“Merck Sharp & Dohme Corp., a subsidiary of Merck & Co., Inc., Kenilworth, NJ, USA, provided financial support for the study.” The authors would like to thank Dr. Edson Moreira for his review and data analysis and Leila Carvalho (employee of MSD Brazil at the time of the study) for her scientific contributions. We acknowledge CTI Clinical Trial & Consulting Services for all editorial support.

## Author Contributions

All authors have read and agreed to the published version of the manuscript and attest that they meet the International Committee of Medical Journal Editors criteria for authorship.

## Conflicts of interest

Fernando B Serra, Paula M Batista and Thais N F Moreira are employees of Merck Sharp & Dohme Corp., a subsidiary of Merck & Co., Inc., Kenilworth, NJ USA, who may own stock and/or hold stock options in Merck & Co., Inc., Kenilworth, NJ, USA. Diogo Ribeiro is an employee of CTI Clinical Trial & Consulting Services. This company received honoraria from the project funder for support in the study planning and manuscript development.

## Notes

### Author Declarations

This study was approved by the Ethics Committee of Universidade Federal de Sao Paulo

