## Supplementary material for "Adult vaccination in Brazil: a cross-sectional survey on physicians’ prescription habits": Supplementary Material.pdf

**Supplementary Table 1** – Correct answers considered for total score calculation (according to the Brazilian Society of Immunization)

| | **MMR** | **Hepatitis A** | **Hepatitis B** | **Hepatitis A and B** | **Tdap or Tdap-IPV** | **Tetanus and diphtheria** | **Varicella** | **Influenza** | **Meningococcal Conjugate ACWY** | **Meningococcal B** | **Yellow Fever** | **PCV13** | **PPV23** | **Zoster** | **Dengue** | **Bivalent HPV** | **Quadrivalent HPV** |
|---|---|---|---|---|---|---|---|---|---|---|---|---|---|---|---|---|---|
| All patients (regardless of age and risk group) | | X[a)] | X[a)] | X[a)] | X[a)] | X[a)] | | X[a)] | | | | | | | | | |
| Adults - 20 to 59 years old | X | X | X | X | X | X | X[c)] | X | X | X | X | X[b)] | X[b)] | X[d)] | X[e)] | X | X |
| Older adults - ≥ 60 years old | X[f)] | X | X | X | X | X | | X | X | | X | X | X | X | | | |
| Patients with chronic liver disease | | X | X | X | | | X[g)] | X | X | X | | X | X | X[j)] | | | |
| Patients with chronic cardiac disease and/or chronic lung disease | | | | | X | | X[g)] | X | | | | X | X | X[j)] | | | |
| Patients with neoplasms or using immunosuppressive drugs | | X | X | X | X | | | X | X | X | | X | X | | | | |
| Patients with chronic kidney disease | | X | X | X | | | X[g)] | X | X | X | | X | X | X[j)] | | | |
| Patients with anatomical and functional asplenia | | X | X | X | | | X[g)] | X | X | X | | X | X | X | | | |
| Patients with rheumatic disease | | X | X | X | | | X[g)] | X | X | X | | X | X | X[j)] | | X | X |
| Patients with primary immunodeficiencies | | X | X | X | | | X[h)] | X | X | X | | X | X | X[j)] | | X | X |
| Patients with HIV/AIDS | X[i)] | X | X | X | X | | X[i)] | X | X | X | X[i)] | X | X | | | X | X |
| Solid organ transplant candidates and recipients | X[k)] | X | X | X | X | | X[k)] | X | X | | X[k)] | X | X | | | | |
| Patients with hematopoietic stem cell transplant | X[l)] | X | X | X | X | | X[l)] | X | X | | X[m)] | X | X | | | | |
| Patients with diabetes mellitus | | | X | | | | X | X | | | | X | X | | | | |
| None of the above | | | | | | | | | | | | | | | X | | |

HIV, Human Immunodeficiency Virus; AIDS, Acquired Immune Deficiency Syndrome; MMR, Measle, Mumps, and Rubella; Tdap,Tetanus, diphtheria, & acellular pertussis; IPV, Inactivated polio vaccine; PCV13, 13-valent pneumococcal conjugate vaccine; PPV23, 23-valent pneumococcal polysaccharide vaccine; HPV, Human papillomavirus.

a) If physician selected '*All patients (regardless of age and risk group)*' the total score was 1 point (maximum score).

b) If this was the only selected item the total score was 0.

For all the cases below, although the vaccines are recommended for a given population, they are not recommended for all patients within that population. Thus, these options were considered as correct when selected by the physicians, but did not deduct points in the score if they were not selected:

c) Indicated for susceptible adults only.

d) May be recommended in patients ≥50 years, at the physician's discretion.

e) This vaccine is not always recommended for adult patients (only seropositive adults).

f) May be recommended in patients ≥60 years in certain situations (such as in cases of outbreaks), at the physician's discretion.

g) Contraindicated in patients with severe immunosuppression.

h) Contraindicated in patients with cellular or cellular and humoral immunity deficiency or with severe immunosuppression.

i) Contraindicated in patients with CD4 <200 cells/mm$^3$ (<15%).

j) Contraindicated in immunosuppressed patients.

k) Contraindicated in the post-transplant (vaccines may be applied only when the patient is immunocompetent).

l) The use in immunocompromised patients should be evaluated by the physician.

m) The indication is dependent on epidemiological risk and the patient's immunological status.

Note: For vaccines recommended for both adult and older adult populations, but for which no special population is specified in the SBIm immunization schedule for special populations, the option “All patients (regardless of age and risk group)” was considered as the correct one. Additionally, for those vaccines recommended for both adults and older adults, a total score of 1 point was attributed if physicians selected this option. This is because the physicians may have interpreted the “target population” solely as the age groups for which vaccines are recommended, and not the specific special populations. This may have overestimated the physicians’ knowledge regarding the vaccines’ target populations. Although the 13-valent pneumococcal conjugate vaccine (PCV13) and the 23-valent pneumococcal vaccine (PPV23) are also recommended in both adults and older adults, the recommendation in adults is valid only in the presence of certain comorbidities. As a result, the option “All patients (regardless of age and risk group)” was not considered correct for these vaccines.

Lastly, there are various instances in the SBIm recommendations where, though a vaccine is recommended for a given special population, the recommendation does not include all patients within that population (e.g. varicella is recommended for patients with HIV/AIDS, but not in those with CD4 <200 cells/ mm3) (1). Specifying all these particularities as response options would lead to an excessively extensive survey, making it impracticable. Alternatively, the methodology described in Supplementary Table 1 footnotes was followed to avoid penalizing the physicians’ knowledge score in this question (such as physicians who did not select a special population because they were aware that the vaccine is not recommended for all patients of that population). Based on this methodology, and in these cases, the option pertaining to the special population (Supplementary Table 1) was considered as correct, but did not deduct points in the score when not selected.

**Supplementary Table 2** – Correct answers for dosage schedule (according to the Brazilian Society of Immunization)

| Vaccine | Dosage schedule |
|---|---|
| MMR | Two doses |
| Hepatitis A | Two doses |
| Hepatitis B | Three doses |
| Hepatitis A e B | Three doses |
| Tdap or Tdap-IPV | Booster dose every 10 years |
| Tetanus and diphtheria | Booster dose every 10 years |
| Varicella | Two doses (NA for older adults) |
| Influenza | Single annual dose |
| Meningococcal Conjugate ACWY | Single dose (the need for additional doses depends on the epidemiological situation) |
| Meningococcal B | Two doses (NA for older adults) |
| Yellow Fever | Single dose |
| PCV13 | Sequential schedule with two vaccines (total of three doses) [NA for adults] |
| PPV23 | Sequential schedule with two vaccines (total of three doses) [NA for adults] |
| Zoster | Single dose |
| Dengue | Three doses (NA for older adults) |
| Bivalent HPV | Three doses (NA for older adults) |
| Quadrivalent HPV | Three doses (NA for older adults) |

MMR, measle, mumps, and rubella; Tdap, adult tetanus-diphtheria-acellular; IPV, Inactivated polio vaccine ; PCV13, 13-valent pneumococcal conjugate vaccine; PPV23, 23-valent pneumococcal polysaccharide vaccine; HPV, human papillomavirus; NA, not applicable.

**Supplementary Table 3** – Correct answers for availability in the Unified Public Health System (according to the Brazilian Society of Immunization)

| Vaccine | Availability in the SUS |
|---|---|
| MMR | Yes, only in adults |
| Hepatitis A | Nor in adults or older adults |
| Hepatitis B | Yes, in adults and older adults |
| Hepatitis A e B | Nor in adults or older adults |
| Tdap or Tdap-IPV | Nor in adults or older adults |
| Tetanus and diphtheria | Yes, in adults and older adults |
| Varicella | Nor in adults or older adults |
| Influenza | Yes, in adults and older adults |
| Meningococcal Conjugate ACWY | Nor in adults or older adults |
| Meningococcal B | Nor in adults or older adults |
| Yellow Fever | Yes, in adults and older adults |
| PCV13 | Nor in adults or older adults |
| PPV23* | Yes, only in older adults |
| Zoster | Nor in adults or older adults |
| Dengue | Nor in adults or older adults |
| Bivalent HPV | Nor in adults or older adults |
| Quadrivalent HPV* | Yes, only in adults |

MMR, measle, mumps, and rubella; Tdap, adult tetanus-diphtheria-acellular; IPV, Inactivated polio vaccine; PCV13, 13-valent pneumococcal conjugate vaccine; PPV23, 23-valent pneumococcal polysaccharide vaccine; HPV, human papillomavirus; NA, not applicable.
*Correct answers were not defined based on SBIm immunization schedules

Note: Vaccines free of charge at the Basic Health Units (BHUs) for a subset of adult and older adult patients were considered as being available in SUS for these age groups. For instance, though PPV23 is free of charge at BHUs for institutionalized older adult patients only, the option "Yes, in older adults" was considered as the correct answer. Additionally, for the Human papillomavirus (HPV) vaccine, the correct answer was not defined based on the SBIm immunization schedules. According to these schedules, no HPV vaccine is free of charge at the BHUs for the adult population. However, a document issued by the Ministry of Health (2) states that the quadrivalent HPV is in fact available at the BHUs for men and women up to 26 years old with HIV/AIDS or immunosuppressed (oncology patients and those submitted to a solid organ or bone marrow transplantation). Thus, the correct answer was considered as "Yes, in adults". For PCV13, the correct answer considered was "Nor in adult or older adults", according to the National Vaccination Calendar at the time of this survey (3).

**Supplementary Table 4 –** Number of required completed surveys per medical specialty and Brazilian region

| Region / Specialty | North | Northeast | Midwest | Southeast | South | Total |
|---|---|---|---|---|---|---|
| Cardiology | 11 | 44 | 20 | 133 | 36 | 244 |
| GP/FM | 4 | 15 | 7 | 46 | 13 | 85 |
| Endocrinology | 4 | 14 | 7 | 43 | 12 | 80 |
| Geriatrics | 1 | 5 | 2 | 14 | 4 | 26 |
| Gynecology | 23 | 92 | 42 | 280 | 78 | 515 |
| Infectious disease | 3 | 10 | 5 | 32 | 9 | 59 |
| Pulmonology | 2 | 11 | 5 | 32 | 9 | 59 |
| Total | 48 | 191 | 88 | 580 | 161 | **1068** |

GP/FP, general practitioners/family medicine

**Supplementary Table 5 –** Proportions of physicians not prescribing the vaccines recommended by the Brazilian Society of Immunization to all adult and older adult patients with an indication

| **Vaccine** | **Adults** | **Older adults** |
|---|---|---|
| MMR | 18.2% (100 out of 550) | 12.4% (31 out of 249) |
| Hepatitis A | 22.0% (82 out of 373) | 14.4% (37 out of 257) |
| Hepatitis B | 10.9% (101 out of 925) | 12.2% (77 out of 630) |
| Hepatitis A and B | 19.8% (99 out of 499) | 11.8% (33 out of 279) |
| Tdap or Tdap-IPV | 12.0% (86 out of 719) | 18.3% (69 out of 378) |
| Tetanus and diphtheria | 14.5% (112 out of 774) | 15.5% (86 out of 556) |
| Varicella | 18.6% (52 out of 279) | NA |
| Influenza | 6.7% (68 out of 1013) | 8.4% (83 out of 991) |
| Meningococcal Conjugate ACWY | 22.7% (49 out of 216) | 14.4% (23 out of 160) |
| Meningococcal B | 18.9% (38 out of 201) | NA |
| Yellow Fever | 12.5% (104 out of 830) | 17.0% (106 out of 625) |
| PCV13 | 16.9% (89 out of 528) | 16.8% (90 out of 536) |
| PPV23 | 12.6% (62 out of 494) | 13.8% (75 out of 544) |
| Zoster | 24.0% (119 out of 496) | 23.1% (120 out of 520) |
| Dengue | 16.4% (35 out of 213) | NA |
| Bivalent HPV | 12.8% (64 out of 499) | NA |
| Quadrivalent HPV | 9.1% (58 out of 638) | NA |

MMR, measle, mumps, and rubella; Tdap, adult tetanus-diphtheria-acellular; IPV,Inactivated polio vaccine; PCV13, 13-valent pneumococcal conjugate vaccine; PPV23, 23-valent pneumococcal polysaccharide vaccine; HPV, human papillomavirus; NA, not applicable.
Note: Proportions were determined based on the number of physicians who indicated to prescribe each vaccine to adults and older adults.

**Supplementary Table 6 –** Reasons for non-prescription of adult recommended vaccines

| Vaccine, n (%) | Vaccine does not belong to the scope of the physicians' specialty | Physician does not provide care for patients in this age group | Vaccine is not available through SUS | High vaccine cost | Fear of side effects | Doubts regarding the vaccine's indication |
|---|---|---|---|---|---|---|
| MMR | 337 (65.1%) | 51 (9.8%) | 12 (2.3%) | 8 (1.5%) | 12 (2.3%) | 84 (16.2%) |
| Hepatitis A | 432 (62.2%) | 38 (5.5%) | 100 (14.4%) | 41 (5.9%) | 18 (2.6%) | 136 (19.6%) |
| Hepatitis B | 112 (78.3%) | 6 (4.2%) | 3 (2.1%) | 2 (1.4%) | 2 (1.4%) | 16 (11.2%) |
| Hepatitis A and B | 297 (52.2%) | 25 (4.4%) | 105 (18.5%) | 46 (8.1%) | 16 (2.8%) | 96 (16.9%) |
| Tdap or Tdap-IPV | 247 (70.8%) | 23 (6.6%) | 19 (5.4%) | 10 (2.9%) | 12 (3.4%) | 57 (16.3%) |
| Tetanus and diphtheria | 206 (70.1%) | 14 (4.8%) | 6 (2.0%) | 5 (1.7%) | 7 (2.4%) | 30 (10.2%) |
| Varicella | 512 (64.9%) | 62 (7.9%) | 69 (8.7%) | 42 (5.3%) | 29 (3.7%) | 121 (15.3%) |
| Influenza | 33 (60.0%) | 3 (5.5%) | 3 (5.5%) | 3 (5.5%) | 2 (3.6%) | 4 (7.3%) |
| Meningococcal Conjugate ACWY | 611 (71.7%) | 60 (7.0%) | 96 (11.3%) | 91 (10.7%) | 34 (4.0%) | 148 (17.4%) |
| Meningococcal B | 622 (71.7%) | 58 (6.7%) | 74 (8.5%) | 84 (9.7%) | 32 (3.7%) | 146 (16.8%) |
| Yellow Fever | 184 (77.3%) | 7 (2.9%) | 5 (2.1%) | 4 (1.7%) | 10 (4.2%) | 17 (7.1%) |
| PCV13 | 383 (70.9%) | 24 (4.4%) | 44 (8.1%) | 36 (6.7%) | 15 (2.8%) | 82 (15.2%) |
| PPV23 | 398 (69.3%) | 20 (3.5%) | 47 (8.2%) | 42 (7.3%) | 12 (2.1%) | 103 (17.9%) |
| Zoster | 309 (54.0%) | 15 (2.6%) | 80 (14.0%) | 67 (11.7%) | 36 (6.3%) | 145 (25.3%) |
| Dengue | 392 (45.8%) | 14 (1.6%) | 126 (14.7%) | 71 (8.3%) | 114 (13.3%) | 205 (24.0%) |
| Bivalent HPV | 302 (53.1%) | 51 (9.0%) | 36 (6.3%) | 25 (4.4%) | 20 (3.5%) | 67 (11.8%) |
| Quadrivalent HPV | 296 (68.8%) | 47 (10.9%) | 41 (9.5%) | 38 (8.8%) | 19 (4.4%) | 73 (17.0%) |

MMR, measle, mumps, and rubella; Tdap, adult tetanus-diphtheria-acellular; IPV,Inactivated polio vaccine ; PCV13, 13-valent pneumococcal conjugate vaccine; PPV23, 23-valent pneumococcal polysaccharide vaccine; HPV, human papillomavirus.

Note: For each reason, the proportion was determined based on the physicians who stated not to prescribe each vaccine to adult patients.

**Supplementary Table 6 –** Reasons for non-prescription of adult recommended vaccines (continuation)

| Vaccine, n (%) | Doubts regarding the vaccine's efficacy | Lack of time during the appointment | Lack of patient interest | Vaccine is not refundable under the list of the National Supplementary Health Agency | Other reason |
|---|---|---|---|---|---|
| MMR | 9 (1.7%) | 46 (8.9%) | 57 (11.0%) | 11 (2.1%) | 101 (19.5%) |
| Hepatitis A | 33 (4.7%) | 68 (9.8%) | 105 (15.1%) | 27 (3.9%) | 117 (16.8%) |
| Hepatitis B | 2 (1.4%) | 14 (9.8%) | 13 (9.1%) | 2 (1.4%) | 20 (14.0%) |
| Hepatitis A and B | 19 (3.3%) | 45 (7.9%) | 58 (10.2%) | 17 (3.0%) | 124 (21.8%) |
| Tdap or Tdap-IPV | 8 (2.3%) | 34 (9.7%) | 37 (10.6%) | 7 (2.0%) | 52 (14.9%) |
| Tetanus and diphtheria | 7 (2.4%) | 32 (10.9%) | 31 (10.5%) | 6 (2.0%) | 56 (19.0%) |
| Varicella | 20 (2.5%) | 56 (7.1%) | 90 (11.4%) | 19 (2.4%) | 123 (15.6%) |
| Influenza | 4 (7.3%) | 4 (7.3%) | 2 (3.6%) | 1 (1.8%) | 13 (23.6%) |
| Meningococcal Conjugate ACWY | 25 (2.9%) | 49 (5.8%) | 74 (8.7%) | 30 (3.5%) | 90 (10.6%) |
| Meningococcal B | 23 (2.7%) | 56 (6.5%) | 83 (9.6%) | 28 (3.2%) | 114 (13.1%) |
| Yellow Fever | 5 (2.1%) | 16 (6.7%) | 18 (7.6%) | 5 (2.1%) | 38 (16.0%) |
| PCV13 | 12 (2.2%) | 35 (6.5%) | 45 (8.3%) | 13 (2.4%) | 71 (13.1%) |
| PPV23 | 19 (3.3%) | 38 (6.6%) | 45 (7.8%) | 13 (2.3%) | 81 (14.1%) |
| Zoster | 46 (8.0%) | 32 (5.6%) | 50 (8.7%) | 15 (2.6%) | 76 (13.3%) |
| Dengue | 200 (23.4%) | 44 (5.1%) | 59 (6.9%) | 27 (3.2%) | 157 (18.4%) |
| Bivalent HPV | 47 (8.3%) | 24 (4.2%) | 32 (5.6%) | 14 (2.5%) | 159 (27.9%) |
| Quadrivalent HPV | 25 (5.8%) | 24 (5.6%) | 28 (6.5%) | 13 (3.0%) | 50 (11.6%) |

MMR, measle, mumps, and rubella; Tdap, adult tetanus-diphtheria-acellular; IPV,Inactivated polio vaccine; PCV13, 13-valent pneumococcal conjugate vaccine; PPV23, 23-valent pneumococcal polysaccharide vaccine; HPV, human papillomavirus.

Note: For each reason, the proportion was determined based on the physicians who stated not to prescribe each vaccine to adult patients.

**Supplementary Table 7 –** Reasons for non-prescription of older adult recommended vaccines

| Vaccine, n (%) | Vaccine does not belong to the scope of the physicians' specialty | Physician does not provide care for patients in this age group | Vaccine is not available through SUS | High vaccine cost | Fear of side effects | Doubts regarding the vaccine's indication |
|---|---|---|---|---|---|---|
| MMR | 484 (59.1%) | 40 (4.9%) | 16 (2.0%) | 10 (1.2%) | 53 (6.5%) | 156 (19.0%) |
| Hepatitis A | 486 (59.9%) | 32 (3.9%) | 60 (7.4%) | 25 (3.1%) | 27 (3.3%) | 131 (16.2%) |
| Hepatitis B | 263 (60.0%) | 22 (5.0%) | 8 (1.8%) | 5 (1.1%) | 21 (4.8%) | 59 (13.5%) |
| Hepatitis A and B | 418 (53.0%) | 27 (3.4%) | 90 (11.4%) | 59 (7.5%) | 24 (3.0%) | 135 (17.1%) |
| Tdap or Tdap-IPV | 421 (61.0%) | 30 (4.3%) | 38 (5.5%) | 21 (3.0%) | 38 (5.5%) | 119 (17.2%) |
| Tetanus and diphtheria | 329 (64.3%) | 25 (4.9%) | 5 (1.0%) | 5 (1.0%) | 24 (4.7%) | 61 (11.9%) |
| Influenza | 47 (61.0%) | 13 (16.9%) | 2 (2.6%) | 2 (2.6%) | 7 (9.1%) | 2 (2.6%) |
| Meningococcal Conjugate ACWY | 587 (64.6%) | 39 (4.3%) | 78 (8.6%) | 76 (8.4%) | 45 (5.0%) | 164 (18.1%) |
| Yellow Fever | 232 (52.4%) | 21 (4.7%) | 4 (0.9%) | 5 (1.1%) | 121 (27.3%) | 45 (10.2%) |
| PCV13 | 351 (66.0%) | 24 (4.5%) | 42 (7.9%) | 31 (5.8%) | 20 (3.8%) | 68 (12.8%) |
| PPV23 | 352 (67.2%) | 21 (4.0%) | 38 (7.3%) | 33 (6.3%) | 20 (3.8%) | 86 (16.4%) |
| Zoster | 315 (57.5%) | 24 (4.4%) | 61 (11.1%) | 58 (10.6%) | 30 (5.5%) | 126 (23.0%) |

MMR, measle, mumps, and rubella; Tdap, adult tetanus-diphtheria-acellular; IPV,Inactivated polio vaccine; PCV13, 13-valent pneumococcal conjugate vaccine; PPV23, 23-valent pneumococcal polysaccharide vaccine; HPV, human papillomavirus.

Note: For each reason, the proportion was determined based on the physicians who stated not to prescribe each vaccine to older adult patients.

**Supplementary Table 7 –** Reasons for non-prescription of older adult recommended vaccines (continuation)

| Vaccine | Doubts regarding the vaccine's efficacy | Lack of time during the appointment | Lack of patient interest | Vaccine is not refundable under the list of the National Supplementary Health Agency | Other reason |
|---|---|---|---|---|---|
| MMR | 23 (2.8%) | 42 (5.1%) | 59 (7.2%) | 12 (1.5%) | 167 (20.4%) |
| Hepatitis A | 32 (3.9%) | 47 (5.8%) | 76 (9.4%) | 14 (1.7%) | 169 (20.8%) |
| Hepatitis B | 11 (2.5%) | 29 (6.6%) | 40 (9.1%) | 4 (0.9%) | 91 (20.8%) |
| Hepatitis A and B | 32 (4.1%) | 51 (6.5%) | 68 (8.6%) | 18 (2.3%) | 176 (22.3%) |
| Tdap or Tdap-IPV | 14 (2.0%) | 42 (6.1%) | 51 (7.4%) | 15 (2.2%) | 127 (18.4%) |
| Tetanus and diphtheria | 11 (2.1%) | 43 (8.4%) | 42 (8.2%) | 4 (0.8%) | 104 (20.3%) |
| Influenza | 2 (2.6%) | 4 (5.2%) | 2 (2.6%) | 1 (1.3%) | 15 (19.5%) |
| Meningococcal Conjugate ACWY | 30 (3.3%) | 46 (5.1%) | 61 (6.7%) | 22 (2.4%) | 136 (15.0%) |
| Yellow Fever | 14 (3.2%) | 20 (4.5%) | 24 (5.4%) | 5 (1.1%) | 76 (17.2%) |
| PCV13 | 18 (3.4%) | 34 (6.4%) | 26 (4.9%) | 12 (2.3%) | 96 (18.0%) |
| PPV23 | 16 (3.1%) | 37 (7.1%) | 27 (5.2%) | 9 (1.7%) | 74 (14.1%) |
| Zoster | 48 (8.8%) | 38 (6.9%) | 34 (6.2%) | 10 (1.8%) | 65 (11.9%) |

MMR, measle, mumps, and rubella; Tdap, adult tetanus-diphtheria-acellular; IPV,Inactivated polio vaccine; PCV13, 13-valent pneumococcal conjugate vaccine; PPV23, 23-valent pneumococcal polysaccharide vaccine; HPV, human papillomavirus.

Note: For each reason, the proportion was determined based on the physicians who stated not to prescribe each vaccine to older adult patients.

**Supplementary Table 8 –** Reasons for underprescription of adult recommended vaccines

| Vaccine, n (%) | Vaccine is not available through SUS | High vaccine cost | Fear of side effects | Doubts regarding the vaccine's indication | Doubts regarding the vaccine's efficacy |
|---|---|---|---|---|---|
| MMR | 18 (22.2%) | 14 (17.3%) | 10 (12.3%) | 18 (22.2%) | 3 (3.7%) |
| Hepatitis A | 38 (56.7%) | 20 (29.9%) | 1 (1.5%) | 3 (4.5%) | 2 (3.0%) |
| Hepatitis B | 9 (10.5%) | 12 (14.0%) | 6 (7.0%) | 18 (20.9%) | 5 (5.8%) |
| Hepatitis A and B | 33 (37.9%) | 25 (28.7%) | 4 (4.6%) | 8 (9.2%) | 3 (3.4%) |
| Tdap or Tdap-IPV | 16 (22.5%) | 10 (14.1%) | 7 (9.9%) | 13 (18.3%) | 5 (7.0%) |
| Tetanus and diphtheria | 3 (3.4%) | 3 (3.4%) | 8 (9.0%) | 11 (12.4%) | 5 (5.6%) |
| Varicella | 16 (45.7%) | 13 (37.1%) | 7 (20.0%) | 3 (8.6%) | 1 (2.9%) |
| Influenza | 4 (7.4%) | 4 (7.4%) | 10 (18.5%) | 11 (20.4%) | 8 (14.8%) |
| Meningococcal Conjugate ACWY | 25 (64.1%) | 24 (61.5%) | 3 (7.7%) | 7 (17.9%) | 1 (2.6%) |
| Meningococcal B | 14 (48.3%) | 17 (58.6%) | 5 (17.2%) | 5 (17.2%) | 1 (3.4%) |
| Yellow Fever | 0 (0.0%) | 0 (0.0%) | 34 (40.0%) | 19 (22.4%) | 3 (3.5%) |
| PCV13 | 29 (37.7%) | 36 (46.8%) | 6 (7.8%) | 18 (23.4%) | 8 (10.4%) |
| PPV23 | 11 (20.8%) | 20 (37.7%) | 9 (17.0%) | 13 (24.5%) | 6 (11.3%) |
| Zoster | 52 (53.6%) | 67 (69.1%) | 9 (9.3%) | 17 (17.5%) | 9 (9.3%) |
| Dengue | 10 (34.5%) | 14 (48.3%) | 9 (31.0%) | 11 (37.9%) | 10 (34.5%) |
| Bivalent HPV | 16 (30.2%) | 31 (58.5%) | 4 (7.5%) | 9 (17.0%) | 3 (5.7%) |
| Quadrivalent HPV | 22 (47.8%) | 34 (73.9%) | 3 (6.5%) | 6 (13.0%) | 1 (2.2%) |

MMR, measle, mumps, and rubella; Tdap, adult tetanus-diphtheria-acellular; IPV,Inactivated polio vaccine; PCV13, 13-valent pneumococcal conjugate vaccine; PPV23, 23-valent pneumococcal polysaccharide vaccine; HPV, human papillomavirus.

Note: For each reason, the proportion was determined based on the physicians who stated to underprescribe each vaccine to adult patients.

**Supplementary Table 8 –** Reasons for underprescription of adult recommended vaccines (continuation)

| Vaccine, n (%) | Lack of time during the appointment | Lack of patient interest | Vaccine is not refundable under the list of the National Supplementary Health Agency | Other reason |
|---|---|---|---|---|
| MMR | 44 (54.3%) | 49 (60.5%) | 12 (14.8%) | 14 (17.3%) |
| Hepatitis A | 18 (26.9%) | 29 (43.3%) | 9 (13.4%) | 9 (13.4%) |
| Hepatitis B | 44 (51.2%) | 30 (34.9%) | 3 (3.5%) | 16 (18.6%) |
| Hepatitis A and B | 30 (34.5%) | 34 (39.1%) | 11 (12.6%) | 17 (19.5%) |
| Tdap or Tdap-IPV | 34 (47.9%) | 28 (39.4%) | 5 (7.0%) | 16 (22.5%) |
| Tetanus and diphtheria | 54 (60.7%) | 34 (38.2%) | 2 (2.2%) | 16 (18.0%) |
| Varicella | 9 (25.7%) | 11 (31.4%) | 3 (8.6%) | 7 (20.0%) |
| Influenza | 26 (48.1%) | 20 (37.0%) | 3 (5.6%) | 12 (22.2%) |
| Meningococcal Conjugate ACWY | 6 (15.4%) | 9 (23.1%) | 9 (23.1%) | 8 (20.5%) |
| Meningococcal B | 3 (10.3%) | 7 (24.1%) | 6 (20.7%) | 3 (10.3%) |
| Yellow Fever | 33 (38.8%) | 24 (28.2%) | 0 (0.0%) | 16 (18.8%) |
| PCV13 | 17 (22.1%) | 21 (27.3%) | 12 (15.6%) | 12 (15.6%) |
| PPV23 | 21 (39.6%) | 14 (26.4%) | 8 (15.1%) | 9 (17.0%) |
| Zoster | 21 (21.6%) | 22 (22.7%) | 23 (23.7%) | 7 (7.2%) |
| Dengue | 8 (27.6%) | 11 (37.9%) | 6 (20.7%) | 2 (6.9%) |
| Bivalent HPV | 9 (17.0%) | 16 (30.2%) | 6 (11.3%) | 11 (20.8%) |
| Quadrivalent HPV | 8 (17.4%) | 11 (23.9%) | 7 (15.2%) | 4 (8.7%) |

MMR, measle, mumps, and rubella; Tdap, adult tetanus-diphtheria-acellular; IPV,Inactivated polio vaccine ; PCV13, 13-valent pneumococcal conjugate vaccine; PPV23, 23-valent pneumococcal polysaccharide vaccine; HPV, human papillomavirus.

Note: For each reason, the proportion was determined based on the physicians who stated to underprescribe each vaccine to adult patients.

**Supplementary Table 9 –** Reasons for underprescription of older adult recommended vaccines

| Vaccine, n (%) | Vaccine is not available through SUS | High vaccine cost | Fear of side effects | Doubts regarding the vaccine's indication | Doubts regarding the vaccine's efficacy |
|---|---|---|---|---|---|
| MMR | 1 (4.5%) | 1 (4.5%) | 4 (18.2%) | 3 (13.6%) | 0 |
| Hepatitis A | 12 (37.5%) | 9 (28.1%) | 3 (9.4%) | 4 (12.5%) | 0 |
| Hepatitis B | 0 | 3 (4.5%) | 5 (7.5%) | 12 (17.9%) | 3 (4.5%) |
| Hepatitis A and B | 9 (36.0%) | 7 (28.0%) | 1 (4.0%) | 3 (12.0%) | 1 (4.0%) |
| Tdap or Tdap-IPV | 9 (16.1%) | 6 (10.7%) | 6 (10.7%) | 5 (8.9%) | 2 (3.6%) |
| Tetanus and diphtheria | 3 (4.2%) | 3 (4.2%) | 4 (5.6%) | 11 (15.3%) | 2 (2.8%) |
| Influenza | 2 (2.7%) | 5 (6.8%) | 9 (12.2%) | 7 (9.5%) | 8 (10.8%) |
| Meningococcal Conjugate ACWY | 9 (52.9%) | 11 (64.7%) | 2 (11.8%) | 1 (5.9%) | 0 |
| Yellow Fever | 0 | 0 | 34 (47.2%) | 11 (15.3%) | 1 (1.4%) |
| PCV13 | 28 (35.4%) | 38 (48.1%) | 8 (10.1%) | 13 (16.5%) | 6 (7.6%) |
| PPV23 | 16 (22.9%) | 29 (41.4%) | 10 (14.3%) | 11 (15.7%) | 8 (11.4%) |
| Zoster | 59 (57.8%) | 72 (70.6%) | 11 (10.8%) | 14 (13.7%) | 9 (8.8%) |

MMR, measle, mumps, and rubella; Tdap, adult tetanus-diphtheria-acellular; IPV,Inactivated polio vaccine ; PCV13, 13-valent pneumococcal conjugate vaccine; PPV23, 23-valent pneumococcal polysaccharide vaccine; HPV, human papillomavirus.

Note: For each reason, the proportion was determined based on the physicians who stated to underprescribe each vaccine to older adult patients.

**Supplementary Table 9 –** Reasons for underprescription of older adult recommended vaccines (continuation)

| Vaccine, n (%) | Lack of time during the appointment | Lack of patient interest | Vaccine is not refundable under the list of the National Supplementary Health Agency | Other reason |
|---|---|---|---|---|
| MMR | 6 (27.3%) | 14 (63.6%) | 1 (4.5%) | 8 (36.4%) |
| Hepatitis A | 8 (25.0%) | 12 (37.5%) | 5 (15.6%) | 7 (21.9%) |
| Hepatitis B | 29 (43.3%) | 30 (44.8%) | 0 | 20 (29.9%) |
| Hepatitis A and B | 8 (32.0%) | 7 (28.0%) | 3 (12.0%) | 6 (24.0%) |
| Tdap or Tdap-IPV | 20 (35.7%) | 24 (42.9%) | 2 (3.6%) | 14 (25.0%) |
| Tetanus and diphtheria | 36 (50.0%) | 24 (33.3%) | 2 (2.8%) | 23 (31.9%) |
| Influenza | 37 (50.0%) | 26 (35.1%) | 2 (2.7%) | 17 (23.0%) |
| Meningococcal Conjugate ACWY | 3 (17.6%) | 8 (47.1%) | 3 (17.6%) | 1 (5.9%) |
| Yellow Fever | 19 (26.4%) | 12 (16.7%) | 0 | 18 (25.0%) |
| PCV13 | 25 (31.6%) | 19 (24.1%) | 11 (13.9%) | 15 (19.0%) |
| PPV23 | 21 (30.0%) | 16 (22.9%) | 11 (15.7%) | 17 (24.3%) |
| Zoster | 27 (26.5%) | 24 (23.5%) | 21 (20.6%) | 10 (9.8%) |

MMR, measle, mumps, and rubella; Tdap, adult tetanus-diphtheria-acellular; IPV,Inactivated polio vaccine ; PCV13, 13-valent pneumococcal conjugate vaccine; PPV23, 23-valent pneumococcal polysaccharide vaccine; HPV, human papillomavirus.

Note: For each reason, the proportion was determined based on the physicians who stated to underprescribe each vaccine to older adult patients.

**Supplementary Table 10** – Knowledge of vaccines – Target populations in accordance with the recommendations of the Brazilian Society of Immunization by medical specialties

| | Cardiology (n=244) | General Practice/Family Practice (n=85) | Endocrinology (n=80) | Geriatrics (n=26) | Gynecology (n=515) | Infectious Diseases (n=59) | Pulmonology (n=59) |
|---|---|---|---|---|---|---|---|
| **MMR (measles, mumps and rubella), n (%)** | | | | | | | |
| All patients (regardless of age and risk group) | 30 (33.7%) | 18 (30.5%) | 5 (16.7%) | 3 (37.5%) | 93 (29.9%) | 10 (18.2%) | 11 (40.7%) |
| Adults - 20 to 59 years old | 40 (44.9%) | 34 (57.6%) | 17 (56.7%) | 4 (50.0%) | 180 (57.9%) | 43 (78.2%) | 15 (55.6%) |
| Older adults - ≥ 60 years old | 12 (13.5%) | 9 (15.3%) | 5 (16.7%) | 1 (12.5%) | 23 (7.4%) | 9 (16.4%) | 3 (11.1%) |
| Patients with chronic liver disease | 6 (6.7%) | 4 (6.8%) | 3 (10.0%) | 1 (12.5%) | 5 (1.6%) | 9 (16.4%) | 0 |
| Patients with chronic cardiac disease and/or chronic lung disease | 10 (11.2%) | 6 (10.2%) | 7 (23.3%) | 2 (25.0%) | 13 (4.2%) | 6 (10.9%) | 4 (14.8%) |
| Patients with neoplasms or using immunosuppressive drugs | 4 (4.5%) | 5 (8.5%) | 3 (10.0%) | 0 | 9 (2.9%) | 2 (3.6%) | 1 (3.7%) |
| Patients with chronic kidney disease | 6 (6.7%) | 5 (8.5%) | 3 (10.0%) | 1 (12.5%) | 11 (3.5%) | 3 (5.5%) | 0 |
| Patients with anatomical and functional asplenia | 4 (4.5%) | 2 (3.4%) | 6 (20.0%) | 0 | 8 (2.6%) | 4 (7.3%) | 0 |
| Patients with rheumatic disease | 6 (6.7%) | 4 (6.8%) | 4 (13.3%) | 1 (12.5%) | 8 (2.6%) | 4 (7.3%) | 1 (3.7%) |
| Patients with primary immunodeficiencies | 5 (5.6%) | 6 (10.2%) | 2 (6.7%) | 1 (12.5%) | 10 (3.2%) | 0 | 1 (3.7%) |
| Patients with HIV/AIDS | 6 (6.7%) | 7 (11.9%) | 5 (16.7%) | 1 (12.5%) | 11 (3.5%) | 11 (20.0%) | 0 |
| Solid organ transplant candidates and recipients | 9 (10.1%) | 4 (6.8%) | 4 (13.3%) | 0 | 11 (3.5%) | 3 (5.5%) | 1 (3.7%) |
| Patients with hematopoietic stem cell transplant | 4 (4.5%) | 2 (3.4%) | 3 (10.0%) | 0 | 5 (1.6%) | 1 (1.8%) | 0 |
| Patients with diabetes mellitus | 10 (11.2%) | 5 (8.5%) | 5 (16.7%) | 0 | 9 (2.9%) | 7 (12.7%) | 1 (3.7%) |
| None of the above | 9 (10.1%) | 3 (5.1%) | 2 (6.7%) | 1 (12.5%) | 35 (11.3%) | 0 | 0 |
| **Total score** | | | | | | | |
| N | 89 | 59 | 30 | 8 | 311 | 55 | 27 |
| Mean | 0.37 | 0.47 | 0.45 | 0.25 | 0.53 | 0.65 | 0.44 |
| Standard deviation | 0.47 | 0.48 | 0.50 | 0.46 | 0.49 | 0.45 | 0.49 |
| Median | 0.00 | 0.33 | 0.00 | 0.00 | 1.00 | 1.00 | 0.00 |
| Minimum | 0.00 | 0.00 | 0.00 | 0.00 | 0.00 | 0.00 | 0.00 |
| Maximum | 1.00 | 1.00 | 1.00 | 1.00 | 1.00 | 1.00 | 1.00 |
| **Hepatitis A, n (%)** | | | | | | | |
| All patients (regardless of age and risk group) | 48 (57.8%) | 20 (42.6%) | 16 (51.6%) | 4 (36.4%) | 73 (48.3%) | 23 (41.8%) | 12 (66.7%) |
| Adults - 20 to 59 years old | 17 (20.5%) | 18 (38.3%) | 6 (19.4%) | 1 (9.1%) | 44 (29.1%) | 14 (25.5%) | 6 (33.3%) |
| Older adults - ≥ 60 years old | 6 (7.2%) | 6 (12.8%) | 1 (3.2%) | 0 | 11 (7.3%) | 6 (10.9%) | 0 |
| Patients with chronic liver disease | 11 (13.3%) | 9 (19.1%) | 8 (25.8%) | 0 | 12 (7.9%) | 25 (45.5%) | 2 (11.1%) |
| Patients with chronic cardiac disease and/or chronic lung disease | 4 (4.8%) | 3 (6.4%) | 4 (12.9%) | 0 | 10 (6.6%) | 3 (5.5%) | 2 (11.1%) |
| Patients with neoplasms or using immunosuppressive drugs | 4 (4.8%) | 1 (2.1%) | 2 (6.5%) | 0 | 10 (6.6%) | 5 (9.1%) | 1 (5.6%) |
| Patients with chronic kidney disease | 4 (4.8%) | 4 (8.5%) | 3 (9.7%) | 0 | 8 (5.3%) | 4 (7.3%) | 1 (5.6%) |

| | | | | | | | |
|---|---|---|---|---|---|---|---|
| Patients with anatomical and functional asplenia | 3 (3.6%) | 0 | 3 (9.7%) | 0 | 7 (4.6%) | 5 (9.1%) | 0 |
| Patients with rheumatic disease | 3 (3.6%) | 3 (6.4%) | 3 (9.7%) | 0 | 7 (4.6%) | 4 (7.3%) | 0 |
| Patients with primary immunodeficiencies | 9 (10.8%) | 1 (2.1%) | 4 (12.9%) | 1 (9.1%) | 11 (7.3%) | 9 (16.4%) | 1 (5.6%) |
| Patients with HIV/AIDS | 11 (13.3%) | 5 (10.6%) | 3 (9.7%) | 1 (9.1%) | 18 (11.9%) | 25 (45.5%) | 0 |
| Solid organ transplant candidates and recipients | 6 (7.2%) | 2 (4.3%) | 4 (12.9%) | 0 | 10 (6.6%) | 12 (21.8%) | 0 |
| Patients with hematopoietic stem cell transplant | 5 (6.0%) | 2 (4.3%) | 2 (6.5%) | 0 | 6 (4.0%) | 6 (10.9%) | 0 |
| Patients with diabetes mellitus | 8 (9.6%) | 5 (10.6%) | 4 (12.9%) | 0 | 9 (6.0%) | 6 (10.9%) | 0 |
| None of the above | 8 (9.6%) | 4 (8.5%) | 2 (6.5%) | 5 (45.5%) | 20 (13.2%) | 0 | 0 |
| **Total score** | | | | | | | |
| N | 83 | 47 | 31 | 11 | 151 | 55 | 18 |
| Mean | 0.65 | 0.51 | 0.61 | 0.39 | 0.56 | 0.59 | 0.71 |
| Standard deviation | 0.43 | 0.44 | 0.43 | 0.49 | 0.45 | 0.38 | 0.42 |
| Median | 1.00 | 0.36 | 1.00 | 0.09 | 0.64 | 0.55 | 1.00 |
| Minimum | 0.00 | 0.00 | 0.00 | 0.00 | 0.00 | 0.09 | 0.09 |
| Maximum | 1.00 | 1.00 | 1.00 | 1.00 | 1.00 | 1.00 | 1.00 |
| **Hepatitis B, n (%)** | | | | | | | |
| All patients (regardless of age and risk group) | 105 (56.1%) | 41 (54.7%) | 39 (60.0%) | 11 (47.8%) | 280 (57.3%) | 42 (71.2%) | 23 (62.2%) |
| Adults - 20 to 59 years old | 51 (27.3%) | 30 (40.0%) | 14 (21.5%) | 6 (26.1%) | 163 (33.3%) | 15 (25.4%) | 12 (32.4%) |
| Older adults - ≥ 60 years old | 26 (13.9%) | 17 (22.7%) | 7 (10.8%) | 6 (26.1%) | 47 (9.6%) | 6 (10.2%) | 3 (8.1%) |
| Patients with chronic liver disease | 22 (11.8%) | 9 (12.0%) | 11 (16.9%) | 5 (21.7%) | 29 (5.9%) | 8 (13.6%) | 9 (24.3%) |
| Patients with chronic cardiac disease and/or chronic lung disease | 19 (10.2%) | 4 (5.3%) | 7 (10.8%) | 5 (21.7%) | 16 (3.3%) | 6 (10.2%) | 6 (16.2%) |
| Patients with neoplasms or using immunosuppressive drugs | 16 (8.6%) | 4 (5.3%) | 5 (7.7%) | 2 (8.7%) | 16 (3.3%) | 3 (5.1%) | 1 (2.7%) |
| Patients with chronic kidney disease | 19 (10.2%) | 6 (8.0%) | 6 (9.2%) | 4 (17.4%) | 19 (3.9%) | 8 (13.6%) | 2 (5.4%) |
| Patients with anatomical and functional asplenia | 12 (6.4%) | 3 (4.0%) | 5 (7.7%) | 3 (13.0%) | 16 (3.3%) | 3 (5.1%) | 4 (10.8%) |
| Patients with rheumatic disease | 7 (3.7%) | 3 (4.0%) | 5 (7.7%) | 2 (8.7%) | 11 (2.2%) | 4 (6.8%) | 0 |
| Patients with primary immunodeficiencies | 19 (10.2%) | 3 (4.0%) | 6 (9.2%) | 1 (4.3%) | 23 (4.7%) | 3 (5.1%) | 3 (8.1%) |
| Patients with HIV/AIDS | 27 (14.4%) | 6 (8.0%) | 9 (13.8%) | 1 (4.3%) | 37 (7.6%) | 7 (11.9%) | 2 (5.4%) |
| Solid organ transplant candidates and recipients | 22 (11.8%) | 4 (5.3%) | 6 (9.2%) | 2 (8.7%) | 28 (5.7%) | 6 (10.2%) | 5 (13.5%) |
| Patients with hematopoietic stem cell transplant | 18 (9.6%) | 3 (4.0%) | 5 (7.7%) | 1 (4.3%) | 14 (2.9%) | 7 (11.9%) | 3 (8.1%) |
| Patients with diabetes mellitus | 16 (8.6%) | 6 (8.0%) | 7 (10.8%) | 3 (13.0%) | 16 (3.3%) | 4 (6.8%) | 2 (5.4%) |
| None of the above | 5 (2.7%) | 0 | 4 (6.2%) | 2 (8.7%) | 31 (6.3%) | 1 (1.7%) | 0 |
| **Total score** | | | | | | | |
| N | 187 | 75 | 65 | 23 | 489 | 59 | 37 |
| Mean | 0.67 | 0.65 | 0.70 | 0.59 | 0.64 | 0.81 | 0.71 |
| Standard deviation | 0.40 | 0.40 | 0.41 | 0.43 | 0.43 | 0.33 | 0.39 |
| Median | 1.00 | 1.00 | 1.00 | 0.83 | 1.00 | 1.00 | 1.00 |
| Minimum | 0.00 | 0.08 | 0.00 | 0.00 | 0.00 | 0.00 | 0.08 |
| Maximum | 1.00 | 1.00 | 1.00 | 1.00 | 1.00 | 1.00 | 1.00 |
| **Hepatitis A and B, n (%)** | | | | | | | |

| | | | | | | | |
|---|---|---|---|---|---|---|---|
| All patients (regardless of age and risk group) | 52 (51.0%) | 18 (33.3%) | 15 (36.6%) | 5 (41.7%) | 111 (45.5%) | 22 (53.7%) | 13 (59.1%) |
| Adults - 20 to 59 years old | 22 (21.6%) | 20 (37.0%) | 14 (34.1%) | 3 (25.0%) | 81 (33.2%) | 13 (31.7%) | 5 (22.7%) |
| Older adults - ≥ 60 years old | 16 (15.7%) | 15 (27.8%) | 2 (4.9%) | 1 (8.3%) | 24 (9.8%) | 3 (7.3%) | 0 |
| Patients with chronic liver disease | 15 (14.7%) | 9 (16.7%) | 14 (34.1%) | 3 (25.0%) | 20 (8.2%) | 15 (36.6%) | 3 (13.6%) |
| Patients with chronic cardiac disease and/or chronic lung disease | 12 (11.8%) | 5 (9.3%) | 7 (17.1%) | 2 (16.7%) | 13 (5.3%) | 3 (7.3%) | 4 (18.2%) |
| Patients with neoplasms or using immunosuppressive drugs | 10 (9.8%) | 4 (7.4%) | 7 (17.1%) | 0 | 14 (5.7%) | 5 (12.2%) | 2 (9.1%) |
| Patients with chronic kidney disease | 10 (9.8%) | 5 (9.3%) | 8 (19.5%) | 2 (16.7%) | 13 (5.3%) | 6 (14.6%) | 1 (4.5%) |
| Patients with anatomical and functional asplenia | 7 (6.9%) | 3 (5.6%) | 8 (19.5%) | 1 (8.3%) | 13 (5.3%) | 3 (7.3%) | 3 (13.6%) |
| Patients with rheumatic disease | 5 (4.9%) | 5 (9.3%) | 7 (17.1%) | 1 (8.3%) | 13 (5.3%) | 4 (9.8%) | 1 (4.5%) |
| Patients with primary immunodeficiencies | 13 (12.7%) | 4 (7.4%) | 6 (14.6%) | 0 | 16 (6.6%) | 6 (14.6%) | 2 (9.1%) |
| Patients with HIV/AIDS | 16 (15.7%) | 4 (7.4%) | 7 (17.1%) | 0 | 21 (8.6%) | 13 (31.7%) | 1 (4.5%) |
| Solid organ transplant candidates and recipients | 12 (11.8%) | 4 (7.4%) | 6 (14.6%) | 0 | 18 (7.4%) | 8 (19.5%) | 1 (4.5%) |
| Patients with hematopoietic stem cell transplant | 7 (6.9%) | 3 (5.6%) | 6 (14.6%) | 0 | 10 (4.1%) | 7 (17.1%) | 1 (4.5%) |
| Patients with diabetes mellitus | 10 (9.8%) | 6 (11.1%) | 5 (12.2%) | 1 (8.3%) | 6 (2.5%) | 3 (7.3%) | 0 |
| None of the above | 14 (13.7%) | 10 (18.5%) | 4 (9.8%) | 2 (16.7%) | 35 (14.3%) | 0 | 2 (9.1%) |
| **Total score** | | | | | | | |
| N | 102 | 54 | 41 | 12 | 244 | 41 | 22 |
| Mean | 0.61 | 0.44 | 0.53 | 0.48 | 0.54 | 0.71 | 0.66 |
| Standard deviation | 0.43 | 0.43 | 0.42 | 0.47 | 0.45 | 0.36 | 0.45 |
| Median | 1.00 | 0.18 | 0.64 | 0.23 | 0.59 | 1.00 | 1.00 |
| Minimum | 0.00 | 0.00 | 0.00 | 0.00 | 0.00 | 0.09 | 0.00 |
| Maximum | 1.00 | 1.00 | 1.00 | 1.00 | 1.00 | 1.00 | 1.00 |
| **Tdap or Tdap-IPV, n (%)** | | | | | | | |
| All patients (regardless of age and risk group) | 49 (45.8%) | 25 (40.3%) | 18 (42.9%) | 7 (35.0%) | 185 (42.0%) | 26 (44.8%) | 15 (46.9%) |
| Adults - 20 to 59 years old | 28 (26.2%) | 18 (29.0%) | 7 (16.7%) | 6 (30.0%) | 147 (33.4%) | 15 (25.9%) | 13 (40.6%) |
| Older adults - ≥ 60 years old | 13 (12.1%) | 13 (21.0%) | 6 (14.3%) | 3 (15.0%) | 40 (9.1%) | 7 (12.1%) | 9 (28.1%) |
| Patients with chronic liver disease | 9 (8.4%) | 3 (4.8%) | 3 (7.1%) | 1 (5.0%) | 16 (3.6%) | 3 (5.2%) | 3 (9.4%) |
| Patients with chronic cardiac disease and/or chronic lung disease | 13 (12.1%) | 2 (3.2%) | 4 (9.5%) | 1 (5.0%) | 24 (5.5%) | 5 (8.6%) | 9 (28.1%) |
| Patients with neoplasms or using immunosuppressive drugs | 4 (3.7%) | 3 (4.8%) | 6 (14.3%) | 1 (5.0%) | 21 (4.8%) | 7 (12.1%) | 2 (6.3%) |
| Patients with chronic kidney disease | 9 (8.4%) | 5 (8.1%) | 4 (9.5%) | 1 (5.0%) | 17 (3.9%) | 4 (6.9%) | 2 (6.3%) |
| Patients with anatomical and functional asplenia | 5 (4.7%) | 4 (6.5%) | 6 (14.3%) | 1 (5.0%) | 18 (4.1%) | 7 (12.1%) | 2 (6.3%) |
| Patients with rheumatic disease | 8 (7.5%) | 3 (4.8%) | 2 (4.8%) | 3 (15.0%) | 14 (3.2%) | 6 (10.3%) | 2 (6.3%) |
| Patients with primary immunodeficiencies | 6 (5.6%) | 7 (11.3%) | 5 (11.9%) | 0 | 28 (6.4%) | 8 (13.8%) | 2 (6.3%) |
| Patients with HIV/AIDS | 8 (7.5%) | 6 (9.7%) | 5 (11.9%) | 2 (10.0%) | 37 (8.4%) | 7 (12.1%) | 2 (6.3%) |
| Solid organ transplant candidates and recipients | 8 (7.5%) | 4 (6.5%) | 6 (14.3%) | 1 (5.0%) | 25 (5.7%) | 9 (15.5%) | 1 (3.1%) |
| Patients with hematopoietic stem cell transplant | 6 (5.6%) | 3 (4.8%) | 5 (11.9%) | 0 | 22 (5.0%) | 9 (15.5%) | 1 (3.1%) |
| Patients with diabetes mellitus | 9 (8.4%) | 5 (8.1%) | 4 (9.5%) | 1 (5.0%) | 16 (3.6%) | 3 (5.2%) | 3 (9.4%) |
| None of the above | 16 (15.0%) | 10 (16.1%) | 7 (16.7%) | 5 (25.0%) | 85 (19.3%) | 6 (10.3%) | 1 (3.1%) |

| | | | | | | | |
|---|---|---|---|---|---|---|---|
| **Total score** | | | | | | | |
| N | 107 | 62 | 42 | 20 | 440 | 58 | 32 |
| Mean | 0.52 | 0.48 | 0.50 | 0.41 | 0.49 | 0.53 | 0.59 |
| Standard deviation | 0.45 | 0.44 | 0.46 | 0.45 | 0.44 | 0.44 | 0.41 |
| Median | 0.29 | 0.29 | 0.29 | 0.14 | 0.29 | 0.36 | 0.57 |
| Minimum | 0.00 | 0.00 | 0.00 | 0.00 | 0.00 | 0.00 | 0.00 |
| Maximum | 1.00 | 1.00 | 1.00 | 1.00 | 1.00 | 1.00 | 1.00 |
| **Tetanus and diphtheria, n (%)** | | | | | | | |
| All patients (regardless of age and risk group) | 80 (56.3%) | 34 (47.9%) | 33 (63.5%) | 13 (61.9%) | 234 (54.4%) | 43 (75.4%) | 20 (60.6%) |
| Adults - 20 to 59 years old | 36 (25.4%) | 29 (40.8%) | 13 (25.0%) | 6 (28.6%) | 151 (35.1%) | 13 (22.8%) | 11 (33.3%) |
| Older adults - ≥ 60 years old | 28 (19.7%) | 18 (25.4%) | 6 (11.5%) | 8 (38.1%) | 52 (12.1%) | 7 (12.3%) | 3 (9.1%) |
| Patients with chronic liver disease | 12 (8.5%) | 3 (4.2%) | 0 | 1 (4.8%) | 7 (1.6%) | 3 (5.3%) | 0 |
| Patients with chronic cardiac disease and/or chronic lung disease | 11 (7.7%) | 4 (5.6%) | 1 (1.9%) | 1 (4.8%) | 7 (1.6%) | 2 (3.5%) | 0 |
| Patients with neoplasms or using immunosuppressive drugs | 2 (1.4%) | 1 (1.4%) | 1 (1.9%) | 0 | 6 (1.4%) | 0 | 0 |
| Patients with chronic kidney disease | 7 (4.9%) | 1 (1.4%) | 0 | 1 (4.8%) | 5 (1.2%) | 2 (3.5%) | 0 |
| Patients with anatomical and functional asplenia | 2 (1.4%) | 1 (1.4%) | 0 | 0 | 4 (0.9%) | 1 (1.8%) | 0 |
| Patients with rheumatic disease | 4 (2.8%) | 0 | 0 | 2 (9.5%) | 2 (0.5%) | 2 (3.5%) | 0 |
| Patients with primary immunodeficiencies | 4 (2.8%) | 2 (2.8%) | 0 | 0 | 7 (1.6%) | 1 (1.8%) | 0 |
| Patients with HIV/AIDS | 9 (6.3%) | 3 (4.2%) | 0 | 1 (4.8%) | 8 (1.9%) | 5 (8.8%) | 0 |
| Solid organ transplant candidates and recipients | 7 (4.9%) | 0 | 0 | 0 | 7 (1.6%) | 3 (5.3%) | 0 |
| Patients with hematopoietic stem cell transplant | 3 (2.1%) | 0 | 0 | 0 | 3 (0.7%) | 1 (1.8%) | 0 |
| Patients with diabetes mellitus | 8 (5.6%) | 4 (5.6%) | 3 (5.8%) | 1 (4.8%) | 7 (1.6%) | 1 (1.8%) | 0 |
| None of the above | 13 (9.2%) | 4 (5.6%) | 3 (5.8%) | 0 | 41 (9.5%) | 1 (1.8%) | 2 (6.1%) |
| **Total score** | | | | | | | |
| N | 142 | 71 | 52 | 21 | 430 | 57 | 33 |
| Mean | 0.71 | 0.73 | 0.81 | 0.88 | 0.75 | 0.86 | 0.82 |
| Standard deviation | 0.41 | 0.39 | 0.36 | 0.31 | 0.36 | 0.31 | 0.30 |
| Median | 1.00 | 1.00 | 1.00 | 1.00 | 1.00 | 1.00 | 1.00 |
| Minimum | 0.00 | 0.00 | 0.00 | 0.00 | 0.00 | 0.00 | 0.00 |
| Maximum | 1.00 | 1.00 | 1.00 | 1.00 | 1.00 | 1.00 | 1.00 |
| **Varicella, n (%)** | | | | | | | |
| All patients (regardless of age and risk group) | 19 (37.3%) | 11 (26.2%) | 6 (40.0%) | 4 (44.4%) | 39 (37.1%) | 8 (18.2%) | 6 (46.2%) |
| Adults - 20 to 59 years old | 18 (35.3%) | 19 (45.2%) | 3 (20.0%) | 2 (22.2%) | 38 (36.2%) | 21 (47.7%) | 4 (30.8%) |
| Older adults - ≥ 60 years old | 10 (19.6%) | 7 (16.7%) | 3 (20.0%) | 0 | 11 (10.5%) | 11 (25.0%) | 4 (30.8%) |
| Patients with chronic liver disease | 6 (11.8%) | 5 (11.9%) | 2 (13.3%) | 0 | 3 (2.9%) | 7 (15.9%) | 0 |
| Patients with chronic cardiac disease and/or chronic lung disease | 9 (17.6%) | 4 (9.5%) | 2 (13.3%) | 0 | 3 (2.9%) | 5 (11.4%) | 1 (7.7%) |
| Patients with neoplasms or using immunosuppressive drugs | 4 (7.8%) | 3 (7.1%) | 2 (13.3%) | 0 | 4 (3.8%) | 4 (9.1%) | 1 (7.7%) |
| Patients with chronic kidney disease | 3 (5.9%) | 3 (7.1%) | 2 (13.3%) | 0 | 4 (3.8%) | 5 (11.4%) | 0 |

| | | | | | | | |
|---|---|---|---|---|---|---|---|
| Patients with anatomical and functional asplenia | 5 (9.8%) | 2 (4.8%) | 3 (20.0%) | 0 | 2 (1.9%) | 3 (6.8%) | 0 |
| Patients with rheumatic disease | 2 (3.9%) | 3 (7.1%) | 2 (13.3%) | 0 | 6 (5.7%) | 5 (11.4%) | 0 |
| Patients with primary immunodeficiencies | 6 (11.8%) | 2 (4.8%) | 3 (20.0%) | 0 | 6 (5.7%) | 5 (11.4%) | 0 |
| Patients with HIV/AIDS | 7 (13.7%) | 2 (4.8%) | 2 (13.3%) | 0 | 6 (5.7%) | 7 (15.9%) | 0 |
| Solid organ transplant candidates and recipients | 4 (7.8%) | 2 (4.8%) | 3 (20.0%) | 0 | 5 (4.8%) | 6 (13.6%) | 0 |
| Patients with hematopoietic stem cell transplant | 2 (3.9%) | 1 (2.4%) | 3 (20.0%) | 0 | 5 (4.8%) | 4 (9.1%) | 0 |
| Patients with diabetes mellitus | 6 (11.8%) | 3 (7.1%) | 2 (13.3%) | 0 | 3 (2.9%) | 8 (18.2%) | 0 |
| None of the above | 4 (7.8%) | 8 (19.0%) | 1 (6.7%) | 3 (33.3%) | 21 (20.0%) | 5 (11.4%) | 2 (15.4%) |
| **Total score** | | | | | | | |
| N | 51 | 42 | 15 | 9 | 105 | 44 | 13 |
| Mean | 0.42 | 0.42 | 0.38 | 0.22 | 0.35 | 0.53 | 0.08 |
| Standard deviation | 0.46 | 0.48 | 0.49 | 0.44 | 0.47 | 0.47 | 0.28 |
| Median | 0.00 | 0.00 | 0.00 | 0.00 | 0.00 | 0.76 | 0.00 |
| Minimum | 0.00 | 0.00 | 0.00 | 0.00 | 0.00 | 0.00 | 0.00 |
| Maximum | 1.00 | 1.00 | 1.00 | 1.00 | 1.00 | 1.00 | 1.00 |
| **Influenza, n (%)** | | | | | | | |
| All patients (regardless of age and risk group) | 96 (40.2%) | 32 (39.0%) | 21 (27.3%) | 10 (38.5%) | 241 (48.9%) | 39 (66.1%) | 21 (36.2%) |
| Adults - 20 to 59 years old | 24 (10.0%) | 15 (18.3%) | 9 (11.7%) | 3 (11.5%) | 75 (15.2%) | 4 (6.8%) | 5 (8.6%) |
| Older adults - ≥ 60 years old | 137 (57.3%) | 44 (53.7%) | 53 (68.8%) | 16 (61.5%) | 225 (45.6%) | 19 (32.2%) | 36 (62.1%) |
| Patients with chronic liver disease | 66 (27.6%) | 20 (24.4%) | 35 (45.5%) | 5 (19.2%) | 80 (16.2%) | 16 (27.1%) | 25 (43.1%) |
| Patients with chronic cardiac disease and/or chronic lung disease | 106 (44.4%) | 32 (39.0%) | 38 (49.4%) | 8 (30.8%) | 119 (24.1%) | 19 (32.2%) | 36 (62.1%) |
| Patients with neoplasms or using immunosuppressive drugs | 53 (22.2%) | 12 (14.6%) | 23 (29.9%) | 3 (11.5%) | 72 (14.6%) | 14 (23.7%) | 24 (41.4%) |
| Patients with chronic kidney disease | 73 (30.5%) | 19 (23.2%) | 32 (41.6%) | 7 (26.9%) | 91 (18.5%) | 18 (30.5%) | 27 (46.6%) |
| Patients with anatomical and functional asplenia | 41 (17.2%) | 10 (12.2%) | 21 (27.3%) | 4 (15.4%) | 57 (11.6%) | 15 (25.4%) | 21 (36.2%) |
| Patients with rheumatic disease | 48 (20.1%) | 16 (19.5%) | 24 (31.2%) | 6 (23.1%) | 74 (15.0%) | 16 (27.1%) | 24 (41.4%) |
| Patients with primary immunodeficiencies | 43 (18.0%) | 15 (18.3%) | 20 (26.0%) | 4 (15.4%) | 86 (17.4%) | 14 (23.7%) | 22 (37.9%) |
| Patients with HIV/AIDS | 51 (21.3%) | 18 (22.0%) | 23 (29.9%) | 5 (19.2%) | 104 (21.1%) | 19 (32.2%) | 25 (43.1%) |
| Solid organ transplant candidates and recipients | 46 (19.2%) | 12 (14.6%) | 18 (23.4%) | 3 (11.5%) | 63 (12.8%) | 16 (27.1%) | 23 (39.7%) |
| Patients with hematopoietic stem cell transplant | 39 (16.3%) | 12 (14.6%) | 16 (20.8%) | 3 (11.5%) | 57 (11.6%) | 14 (23.7%) | 21 (36.2%) |
| Patients with diabetes mellitus | 80 (33.5%) | 28 (34.1%) | 45 (58.4%) | 8 (30.8%) | 104 (21.1%) | 18 (30.5%) | 30 (51.7%) |
| None of the above | 1 (0.4%) | 2 (2.4%) | 0 | 0 | 11 (2.2%) | 0 | 0 |
| **Total score** | | | | | | | |
| N | 239 | 82 | 77 | 26 | 493 | 59 | 58 |
| Mean | 0.66 | 0.63 | 0.63 | 0.61 | 0.68 | 0.92 | 0.79 |
| Standard deviation | 0.37 | 0.38 | 0.35 | 0.41 | 0.39 | 0.16 | 0.29 |
| Median | 0.85 | 0.77 | 0.69 | 0.81 | 0.92 | 1.00 | 0.92 |
| Minimum | 0.00 | 0.00 | 0.08 | 0.08 | 0.00 | 0.08 | 0.15 |
| Maximum | 1.00 | 1.00 | 1.00 | 1.00 | 1.00 | 1.00 | 1.00 |
| **Meningococcal Conjugate ACWY, n (%)** | | | | | | | |

| | | | | | | | |
|---|---|---|---|---|---|---|---|
| All patients (regardless of age and risk group) | 20 (42.6%) | 13 (31.7%) | 5 (38.5%) | 1 (25.0%) | 24 (31.6%) | 20 (44.4%) | 10 (62.5%) |
| Adults - 20 to 59 years old | 11 (23.4%) | 9 (22.0%) | 2 (15.4%) | 2 (50.0%) | 31 (40.8%) | 9 (20.0%) | 5 (31.3%) |
| Older adults - ≥ 60 years old | 10 (21.3%) | 10 (24.4%) | 2 (15.4%) | 0 | 16 (21.1%) | 5 (11.1%) | 3 (18.8%) |
| Patients with chronic liver disease | 5 (10.6%) | 7 (17.1%) | 3 (23.1%) | 0 | 3 (3.9%) | 7 (15.6%) | 0 |
| Patients with chronic cardiac disease and/or chronic lung disease | 12 (25.5%) | 9 (22.0%) | 4 (30.8%) | 0 | 5 (6.6%) | 8 (17.8%) | 3 (18.8%) |
| Patients with neoplasms or using immunosuppressive drugs | 3 (6.4%) | 7 (17.1%) | 2 (15.4%) | 0 | 7 (9.2%) | 7 (15.6%) | 1 (6.3%) |
| Patients with chronic kidney disease | 6 (12.8%) | 6 (14.6%) | 2 (15.4%) | 0 | 4 (5.3%) | 8 (17.8%) | 1 (6.3%) |
| Patients with anatomical and functional asplenia | 8 (17.0%) | 5 (12.2%) | 1 (7.7%) | 0 | 4 (5.3%) | 14 (31.1%) | 0 |
| Patients with rheumatic disease | 4 (8.5%) | 5 (12.2%) | 2 (15.4%) | 0 | 2 (2.6%) | 10 (22.2%) | 0 |
| Patients with primary immunodeficiencies | 7 (14.9%) | 8 (19.5%) | 3 (23.1%) | 0 | 7 (9.2%) | 12 (26.7%) | 1 (6.3%) |
| Patients with HIV/AIDS | 5 (10.6%) | 8 (19.5%) | 1 (7.7%) | 0 | 9 (11.8%) | 16 (35.6%) | 0 |
| Solid organ transplant candidates and recipients | 4 (8.5%) | 5 (12.2%) | 2 (15.4%) | 0 | 5 (6.6%) | 12 (26.7%) | 0 |
| Patients with hematopoietic stem cell transplant | 3 (6.4%) | 6 (14.6%) | 2 (15.4%) | 0 | 5 (6.6%) | 12 (26.7%) | 0 |
| Patients with diabetes mellitus | 7 (14.9%) | 5 (12.2%) | 2 (15.4%) | 0 | 1 (1.3%) | 5 (11.1%) | 0 |
| None of the above | 3 (6.4%) | 6 (14.6%) | 2 (15.4%) | 1 (25.0%) | 13 (17.1%) | 4 (8.9%) | 0 |
| **Total score** | | | | | | | |
| N | 47 | 41 | 13 | 4 | 76 | 45 | 16 |
| Mean | 0.09 | 0.14 | 0.11 | 0.05 | 0.10 | 0.20 | 0.05 |
| Standard deviation | 0.16 | 0.22 | 0.23 | 0.05 | 0.16 | 0.28 | 0.08 |
| Median | 0.00 | 0.00 | 0.00 | 0.05 | 0.09 | 0.00 | 0.00 |
| Minimum | 0.00 | 0.00 | 0.00 | 0.00 | 0.00 | 0.00 | 0.00 |
| Maximum | 0.73 | 0.73 | 0.82 | 0.09 | 0.73 | 0.82 | 0.27 |
| **Meningococcal B, n (%)** | | | | | | | |
| All patients (regardless of age and risk group) | 22 (48.9%) | 17 (43.6%) | 4 (33.3%) | 0 | 21 (38.9%) | 14 (40.0%) | 8 (66.7%) |
| Adults - 20 to 59 years old | 12 (26.7%) | 9 (23.1%) | 1 (8.3%) | 2 (50.0%) | 19 (35.2%) | 12 (34.3%) | 2 (16.7%) |
| Older adults - ≥ 60 years old | 10 (22.2%) | 6 (15.4%) | 1 (8.3%) | 1 (25.0%) | 7 (13.0%) | 5 (14.3%) | 1 (8.3%) |
| Patients with chronic liver disease | 4 (8.9%) | 4 (10.3%) | 2 (16.7%) | 1 (25.0%) | 3 (5.6%) | 6 (17.1%) | 1 (8.3%) |
| Patients with chronic cardiac disease and/or chronic lung disease | 9 (20.0%) | 4 (10.3%) | 2 (16.7%) | 1 (25.0%) | 3 (5.6%) | 7 (20.0%) | 1 (8.3%) |
| Patients with neoplasms or using immunosuppressive drugs | 5 (11.1%) | 3 (7.7%) | 1 (8.3%) | 0 | 3 (5.6%) | 6 (17.1%) | 0 |
| Patients with chronic kidney disease | 4 (8.9%) | 5 (12.8%) | 1 (8.3%) | 1 (25.0%) | 2 (3.7%) | 7 (20.0%) | 0 |
| Patients with anatomical and functional asplenia | 6 (13.3%) | 5 (12.8%) | 0 | 1 (25.0%) | 2 (3.7%) | 9 (25.7%) | 0 |
| Patients with rheumatic disease | 1 (2.2%) | 3 (7.7%) | 1 (8.3%) | 0 | 2 (3.7%) | 7 (20.0%) | 0 |
| Patients with primary immunodeficiencies | 3 (6.7%) | 6 (15.4%) | 1 (8.3%) | 0 | 4 (7.4%) | 8 (22.9%) | 0 |
| Patients with HIV/AIDS | 3 (6.7%) | 6 (15.4%) | 1 (8.3%) | 1 (25.0%) | 7 (13.0%) | 8 (22.9%) | 0 |
| Solid organ transplant candidates and recipients | 3 (6.7%) | 2 (5.1%) | 0 | 1 (25.0%) | 3 (5.6%) | 8 (22.9%) | 0 |
| Patients with hematopoietic stem cell transplant | 3 (6.7%) | 2 (5.1%) | 0 | 0 | 2 (3.7%) | 6 (17.1%) | 0 |
| Patients with diabetes mellitus | 4 (8.9%) | 4 (10.3%) | 2 (16.7%) | 1 (25.0%) | 0 | 3 (8.6%) | 0 |
| None of the above | 2 (4.4%) | 4 (10.3%) | 3 (25.0%) | 1 (25.0%) | 9 (16.7%) | 2 (5.7%) | 2 (16.7%) |

| | | | | | | | |
|---|---|---|---|---|---|---|---|
| **Total score** | | | | | | | |
| N | 45 | 39 | 12 | 4 | 54 | 35 | 12 |
| Mean | 0.06 | 0.08 | 0.03 | 0.10 | 0.07 | 0.18 | 0.01 |
| Standard deviation | 0.14 | 0.17 | 0.07 | 0.14 | 0.16 | 0.27 | 0.03 |
| Median | 0.00 | 0.00 | 0.00 | 0.05 | 0.00 | 0.00 | 0.00 |
| Minimum | 0.00 | 0.00 | 0.00 | 0.00 | 0.00 | 0.00 | 0.00 |
| Maximum | 0.70 | 0.70 | 0.20 | 0.30 | 0.80 | 0.80 | 0.10 |
| **Yellow Fever, n (%)** | | | | | | | |
| All patients (regardless of age and risk group) | 66 (31.3%) | 26 (33.3%) | 14 (21.5%) | 3 (12.5%) | 163 (44.2%) | 11 (18.6%) | 11 (25.6%) |
| Adults - 20 to 59 years old | 91 (43.1%) | 39 (50.0%) | 32 (49.2%) | 9 (37.5%) | 130 (35.2%) | 41 (69.5%) | 20 (46.5%) |
| Older adults - ≥ 60 years old | 31 (14.7%) | 20 (25.6%) | 9 (13.8%) | 2 (8.3%) | 39 (10.6%) | 11 (18.6%) | 9 (20.9%) |
| Patients with chronic liver disease | 9 (4.3%) | 6 (7.7%) | 2 (3.1%) | 0 | 7 (1.9%) | 5 (8.5%) | 2 (4.7%) |
| Patients with chronic cardiac disease and/or chronic lung disease | 19 (9.0%) | 8 (10.3%) | 2 (3.1%) | 1 (4.2%) | 13 (3.5%) | 6 (10.2%) | 3 (7.0%) |
| Patients with neoplasms or using immunosuppressive drugs | 4 (1.9%) | 4 (5.1%) | 0 | 0 | 2 (0.5%) | 1 (1.7%) | 0 |
| Patients with chronic kidney disease | 10 (4.7%) | 6 (7.7%) | 1 (1.5%) | 2 (8.3%) | 7 (1.9%) | 5 (8.5%) | 2 (4.7%) |
| Patients with anatomical and functional asplenia | 6 (2.8%) | 1 (1.3%) | 2 (3.1%) | 0 | 4 (1.1%) | 1 (1.7%) | 1 (2.3%) |
| Patients with rheumatic disease | 10 (4.7%) | 3 (3.8%) | 2 (3.1%) | 1 (4.2%) | 8 (2.2%) | 3 (5.1%) | 2 (4.7%) |
| Patients with primary immunodeficiencies | 3 (1.4%) | 1 (1.3%) | 0 | 0 | 3 (0.8%) | 0 | 0 |
| Patients with HIV/AIDS | 4 (1.9%) | 4 (5.1%) | 1 (1.5%) | 0 | 3 (0.8%) | 6 (10.2%) | 0 |
| Solid organ transplant candidates and recipients | 6 (2.8%) | 0 | 1 (1.5%) | 1 (4.2%) | 1 (0.3%) | 2 (3.4%) | 1 (2.3%) |
| Patients with hematopoietic stem cell transplant | 3 (1.4%) | 1 (1.3%) | 0 | 0 | 1 (0.3%) | 0 | 0 |
| Patients with diabetes mellitus | 24 (11.4%) | 12 (15.4%) | 5 (7.7%) | 1 (4.2%) | 17 (4.6%) | 8 (13.6%) | 3 (7.0%) |
| None of the above | 46 (21.8%) | 10 (12.8%) | 18 (27.7%) | 11 (45.8%) | 70 (19.0%) | 7 (11.9%) | 11 (25.6%) |
| **Total score** | | | | | | | |
| N | 211 | 78 | 65 | 24 | 369 | 59 | 43 |
| Mean | 0.19 | 0.27 | 0.25 | 0.22 | 0.19 | 0.32 | 0.27 |
| Standard deviation | 0.28 | 0.35 | 0.32 | 0.30 | 0.30 | 0.30 | 0.35 |
| Median | 0.00 | 0.00 | 0.00 | 0.00 | 0.00 | 0.50 | 0.00 |
| Minimum | 0.00 | 0.00 | 0.00 | 0.00 | 0.00 | 0.00 | 0.00 |
| Maximum | 1.00 | 1.00 | 1.00 | 1.00 | 1.00 | 1.00 | 1.00 |
| **PCV13, n (%)** | | | | | | | |
| All patients (regardless of age and risk group) | 46 (22.1%) | 18 (26.5%) | 10 (22.7%) | 4 (16.0%) | 46 (35.1%) | 16 (34.0%) | 13 (22.8%) |
| Adults - 20 to 59 years old | 34 (16.3%) | 15 (22.1%) | 3 (6.8%) | 5 (20.0%) | 26 (19.8%) | 3 (6.4%) | 9 (15.8%) |
| Older adults - ≥ 60 years old | 128 (61.5%) | 39 (57.4%) | 26 (59.1%) | 17 (68.0%) | 65 (49.6%) | 22 (46.8%) | 38 (66.7%) |
| Patients with chronic liver disease | 48 (23.1%) | 13 (19.1%) | 10 (22.7%) | 6 (24.0%) | 15 (11.5%) | 12 (25.5%) | 21 (36.8%) |
| Patients with chronic cardiac disease and/or chronic lung disease | 92 (44.2%) | 24 (35.3%) | 19 (43.2%) | 13 (52.0%) | 26 (19.8%) | 19 (40.4%) | 40 (70.2%) |
| Patients with neoplasms or using immunosuppressive drugs | 36 (17.3%) | 11 (16.2%) | 13 (29.5%) | 7 (28.0%) | 17 (13.0%) | 15 (31.9%) | 17 (29.8%) |
| Patients with chronic kidney disease | 49 (23.6%) | 12 (17.6%) | 12 (27.3%) | 8 (32.0%) | 11 (8.4%) | 14 (29.8%) | 23 (40.4%) |

| | | | | | | | |
|---|---|---|---|---|---|---|---|
| Patients with anatomical and functional asplenia | 40 (19.2%) | 12 (17.6%) | 12 (27.3%) | 7 (28.0%) | 9 (6.9%) | 17 (36.2%) | 20 (35.1%) |
| Patients with rheumatic disease | 35 (16.8%) | 13 (19.1%) | 6 (13.6%) | 7 (28.0%) | 10 (7.6%) | 9 (19.1%) | 20 (35.1%) |
| Patients with primary immunodeficiencies | 36 (17.3%) | 14 (20.6%) | 13 (29.5%) | 4 (16.0%) | 16 (12.2%) | 15 (31.9%) | 17 (29.8%) |
| Patients with HIV/AIDS | 42 (20.2%) | 15 (22.1%) | 12 (27.3%) | 5 (20.0%) | 22 (16.8%) | 20 (42.6%) | 18 (31.6%) |
| Solid organ transplant candidates and recipients | 38 (18.3%) | 14 (20.6%) | 10 (22.7%) | 4 (16.0%) | 14 (10.7%) | 14 (29.8%) | 18 (31.6%) |
| Patients with hematopoietic stem cell transplant | 33 (15.9%) | 10 (14.7%) | 9 (20.5%) | 2 (8.0%) | 13 (9.9%) | 13 (27.7%) | 16 (28.1%) |
| Patients with diabetes mellitus | 42 (20.2%) | 15 (22.1%) | 16 (36.4%) | 7 (28.0%) | 11 (8.4%) | 9 (19.1%) | 24 (42.1%) |
| None of the above | 8 (3.8%) | 2 (2.9%) | 4 (9.1%) | 2 (8.0%) | 11 (8.4%) | 4 (8.5%) | 3 (5.3%) |
| **Total score** | | | | | | | |
| N | 208 | 68 | 44 | 25 | 131 | 47 | 57 |
| Mean | 0.24 | 0.23 | 0.28 | 0.28 | 0.15 | 0.29 | 0.38 |
| Standard deviation | 0.28 | 0.29 | 0.31 | 0.31 | 0.24 | 0.35 | 0.36 |
| Median | 0.15 | 0.08 | 0.19 | 0.15 | 0.08 | 0.08 | 0.23 |
| Minimum | 0.00 | 0.00 | 0.00 | 0.00 | 0.00 | 0.00 | 0.00 |
| Maximum | 1.00 | 1.00 | 1.00 | 0.92 | 0.92 | 0.92 | 0.92 |
| **PPV23, n (%)** | | | | | | | |
| All patients (regardless of age and risk group) | 44 (22.6%) | 15 (22.7%) | 9 (24.3%) | 4 (15.4%) | 46 (36.2%) | 11 (19.0%) | 13 (22.4%) |
| Adults - 20 to 59 years old | 27 (13.8%) | 17 (25.8%) | 2 (5.4%) | 3 (11.5%) | 26 (20.5%) | 10 (17.2%) | 11 (19.0%) |
| Older adults - ≥ 60 years old | 123 (63.1%) | 39 (59.1%) | 22 (59.5%) | 18 (69.2%) | 60 (47.2%) | 40 (69.0%) | 40 (69.0%) |
| Patients with chronic liver disease | 40 (20.5%) | 11 (16.7%) | 10 (27.0%) | 10 (38.5%) | 13 (10.2%) | 23 (39.7%) | 22 (37.9%) |
| Patients with chronic cardiac disease and/or chronic lung disease | 91 (46.7%) | 26 (39.4%) | 16 (43.2%) | 15 (57.7%) | 29 (22.8%) | 33 (56.9%) | 41 (70.7%) |
| Patients with neoplasms or using immunosuppressive drugs | 37 (19.0%) | 11 (16.7%) | 11 (29.7%) | 7 (26.9%) | 16 (12.6%) | 23 (39.7%) | 16 (27.6%) |
| Patients with chronic kidney disease | 42 (21.5%) | 14 (21.2%) | 11 (29.7%) | 8 (30.8%) | 11 (8.7%) | 26 (44.8%) | 22 (37.9%) |
| Patients with anatomical and functional asplenia | 36 (18.5%) | 12 (18.2%) | 14 (37.8%) | 9 (34.6%) | 9 (7.1%) | 32 (55.2%) | 19 (32.8%) |
| Patients with rheumatic disease | 34 (17.4%) | 11 (16.7%) | 7 (18.9%) | 7 (26.9%) | 9 (7.1%) | 20 (34.5%) | 18 (31.0%) |
| Patients with primary immunodeficiencies | 33 (16.9%) | 12 (18.2%) | 10 (27.0%) | 5 (19.2%) | 13 (10.2%) | 30 (51.7%) | 15 (25.9%) |
| Patients with HIV/AIDS | 38 (19.5%) | 16 (24.2%) | 10 (27.0%) | 5 (19.2%) | 18 (14.2%) | 35 (60.3%) | 17 (29.3%) |
| Solid organ transplant candidates and recipients | 31 (15.9%) | 11 (16.7%) | 9 (24.3%) | 5 (19.2%) | 17 (13.4%) | 25 (43.1%) | 15 (25.9%) |
| Patients with hematopoietic stem cell transplant | 25 (12.8%) | 9 (13.6%) | 9 (24.3%) | 4 (15.4%) | 10 (7.9%) | 23 (39.7%) | 18 (31.0%) |
| Patients with diabetes mellitus | 40 (20.5%) | 14 (21.2%) | 15 (40.5%) | 11 (42.3%) | 13 (10.2%) | 22 (37.9%) | 22 (37.9%) |
| None of the above | 5 (2.6%) | 1 (1.5%) | 3 (8.1%) | 0 | 11 (8.7%) | 2 (3.4%) | 0 |
| **Total score** | | | | | | | |
| N | 195 | 66 | 37 | 26 | 127 | 58 | 58 |
| Mean | 0.23 | 0.23 | 0.30 | 0.32 | 0.14 | 0.45 | 0.37 |
| Standard deviation | 0.28 | 0.29 | 0.34 | 0.31 | 0.23 | 0.38 | 0.36 |
| Median | 0.15 | 0.12 | 0.15 | 0.23 | 0.08 | 0.42 | 0.23 |
| Minimum | 0.00 | 0.00 | 0.00 | 0.00 | 0.00 | 0.00 | 0.00 |
| Maximum | 1.00 | 0.92 | 1.00 | 0.92 | 0.92 | 1.00 | 1.00 |
| **Zoster, n (%)** | | | | | | | |

| | | | | | | | |
|---|---|---|---|---|---|---|---|
| All patients (regardless of age and risk group) | 22 (24.7%) | 9 (17.6%) | 2 (6.5%) | 2 (8.7%) | 24 (7.8%) | 4 (9.8%) | 6 (31.6%) |
| Adults - 20 to 59 years old | 19 (21.3%) | 18 (35.3%) | 3 (9.7%) | 1 (4.3%) | 88 (28.6%) | 7 (17.1%) | 4 (21.1%) |
| Older adults - ≥ 60 years old | 51 (57.3%) | 26 (51.0%) | 24 (77.4%) | 16 (69.6%) | 222 (72.1%) | 32 (78.0%) | 11 (57.9%) |
| Patients with chronic liver disease | 8 (9.0%) | 2 (3.9%) | 4 (12.9%) | 4 (17.4%) | 6 (1.9%) | 5 (12.2%) | 1 (5.3%) |
| Patients with chronic cardiac disease and/or chronic lung disease | 16 (18.0%) | 2 (3.9%) | 5 (16.1%) | 3 (13.0%) | 30 (9.7%) | 2 (4.9%) | 1 (5.3%) |
| Patients with neoplasms or using immunosuppressive drugs | 8 (9.0%) | 2 (3.9%) | 6 (19.4%) | 3 (13.0%) | 43 (14.0%) | 1 (2.4%) | 1 (5.3%) |
| Patients with chronic kidney disease | 7 (7.9%) | 2 (3.9%) | 3 (9.7%) | 5 (21.7%) | 13 (4.2%) | 3 (7.3%) | 1 (5.3%) |
| Patients with anatomical and functional asplenia | 3 (3.4%) | 3 (5.9%) | 5 (16.1%) | 3 (13.0%) | 9 (2.9%) | 2 (4.9%) | 1 (5.3%) |
| Patients with rheumatic disease | 9 (10.1%) | 2 (3.9%) | 5 (16.1%) | 5 (21.7%) | 21 (6.8%) | 3 (7.3%) | 1 (5.3%) |
| Patients with primary immunodeficiencies | 11 (12.4%) | 2 (3.9%) | 6 (19.4%) | 4 (17.4%) | 40 (13.0%) | 1 (2.4%) | 1 (5.3%) |
| Patients with HIV/AIDS | 9 (10.1%) | 4 (7.8%) | 6 (19.4%) | 4 (17.4%) | 48 (15.6%) | 1 (2.4%) | 1 (5.3%) |
| Solid organ transplant candidates and recipients | 8 (9.0%) | 5 (9.8%) | 6 (19.4%) | 1 (4.3%) | 17 (5.5%) | 2 (4.9%) | 1 (5.3%) |
| Patients with hematopoietic stem cell transplant | 6 (6.7%) | 2 (3.9%) | 5 (16.1%) | 2 (8.7%) | 14 (4.5%) | 1 (2.4%) | 1 (5.3%) |
| Patients with diabetes mellitus | 12 (13.5%) | 4 (7.8%) | 9 (29.0%) | 3 (13.0%) | 37 (12.0%) | 5 (12.2%) | 1 (5.3%) |
| None of the above | 7 (7.9%) | 2 (3.9%) | 2 (6.5%) | 4 (17.4%) | 26 (8.4%) | 3 (7.3%) | 2 (10.5%) |
| **Total score** | | | | | | | |
| N | 89 | 51 | 31 | 23 | 308 | 41 | 19 |
| Mean | 0.29 | 0.29 | 0.31 | 0.36 | 0.33 | 0.40 | 0.31 |
| Standard deviation | 0.28 | 0.25 | 0.26 | 0.28 | 0.24 | 0.21 | 0.29 |
| Median | 0.25 | 0.29 | 0.50 | 0.50 | 0.50 | 0.50 | 0.50 |
| Minimum | 0.00 | 0.00 | 0.00 | 0.00 | 0.00 | 0.00 | 0.00 |
| Maximum | 1.00 | 0.80 | 0.75 | 0.71 | 0.75 | 0.71 | 0.67 |
| **Dengue, n (%)** | | | | | | | |
| All patients (regardless of age and risk group) | 37 (67.3%) | 17 (56.7%) | 7 (58.3%) | 2 (66.7%) | 48 (51.6%) | 3 (25.0%) | 6 (75.0%) |
| Adults - 20 to 59 years old | 12 (21.8%) | 12 (40.0%) | 2 (16.7%) | 1 (33.3%) | 22 (23.7%) | 4 (33.3%) | 2 (25.0%) |
| Older adults - ≥ 60 years old | 4 (7.3%) | 7 (23.3%) | 2 (16.7%) | 0 | 9 (9.7%) | 0 | 0 |
| Patients with chronic liver disease | 1 (1.8%) | 2 (6.7%) | 1 (8.3%) | 0 | 3 (3.2%) | 0 | 0 |
| Patients with chronic cardiac disease and/or chronic lung disease | 1 (1.8%) | 2 (6.7%) | 1 (8.3%) | 0 | 3 (3.2%) | 0 | 0 |
| Patients with neoplasms or using immunosuppressive drugs | 0 | 0 | 0 | 0 | 1 (1.1%) | 0 | 0 |
| Patients with chronic kidney disease | 0 | 1 (3.3%) | 0 | 0 | 1 (1.1%) | 0 | 0 |
| Patients with anatomical and functional asplenia | 1 (1.8%) | 1 (3.3%) | 0 | 0 | 1 (1.1%) | 0 | 0 |
| Patients with rheumatic disease | 0 | 1 (3.3%) | 0 | 0 | 1 (1.1%) | 0 | 0 |
| Patients with primary immunodeficiencies | 0 | 0 | 0 | 0 | 0 | 0 | 0 |
| Patients with HIV/AIDS | 0 | 0 | 1 (8.3%) | 0 | 1 (1.1%) | 0 | 0 |
| Solid organ transplant candidates and recipients | 0 | 0 | 0 | 0 | 0 | 0 | 0 |
| Patients with hematopoietic stem cell transplant | 0 | 0 | 0 | 0 | 0 | 0 | 0 |
| Patients with diabetes mellitus | 3 (5.5%) | 4 (13.3%) | 1 (8.3%) | 0 | 2 (2.2%) | 0 | 0 |
| None of the above | 5 (9.1%) | 1 (3.3%) | 3 (25.0%) | 0 | 20 (21.5%) | 5 (41.7%) | 0 |

| | | | | | | | |
|---|---|---|---|---|---|---|---|
| **Total score** | | | | | | | |
| N | 55 | 30 | 12 | 3 | 93 | 12 | 8 |
| Mean | 0.22 | 0.17 | 0.25 | 0.33 | 0.38 | 0.75 | 0.25 |
| Standard deviation | 0.42 | 0.38 | 0.45 | 0.58 | 0.49 | 0.45 | 0.46 |
| Median | 0.00 | 0.00 | 0.00 | 0.00 | 0.00 | 1.00 | 0.00 |
| Minimum | 0.00 | 0.00 | 0.00 | 0.00 | 0.00 | 0.00 | 0.00 |
| Maximum | 1.00 | 1.00 | 1.00 | 1.00 | 1.00 | 1.00 | 1.00 |
| **Bivalent HPV, n (%)** | | | | | | | |
| All patients (regardless of age and risk group) | 15 (39.5%) | 8 (18.2%) | 2 (9.5%) | 1 (33.3%) | 56 (16.1%) | 7 (20.6%) | 3 (27.3%) |
| Adults - 20 to 59 years old | 11 (28.9%) | 18 (40.9%) | 7 (33.3%) | 2 (66.7%) | 150 (43.1%) | 12 (35.3%) | 4 (36.4%) |
| Older adults - ≥ 60 years old | 1 (2.6%) | 3 (6.8%) | 1 (4.8%) | 0 | 33 (9.5%) | 1 (2.9%) | 0 |
| Patients with chronic liver disease | 1 (2.6%) | 0 | 1 (4.8%) | 0 | 4 (1.1%) | 0 | 0 |
| Patients with chronic cardiac disease and/or chronic lung disease | 2 (5.3%) | 0 | 0 | 0 | 2 (0.6%) | 0 | 1 (9.1%) |
| Patients with neoplasms or using immunosuppressive drugs | 1 (2.6%) | 2 (4.5%) | 0 | 0 | 9 (2.6%) | 0 | 0 |
| Patients with chronic kidney disease | 0 | 0 | 0 | 0 | 6 (1.7%) | 0 | 0 |
| Patients with anatomical and functional asplenia | 1 (2.6%) | 1 (2.3%) | 0 | 0 | 3 (0.9%) | 0 | 0 |
| Patients with rheumatic disease | 1 (2.6%) | 1 (2.3%) | 1 (4.8%) | 0 | 9 (2.6%) | 0 | 0 |
| Patients with primary immunodeficiencies | 2 (5.3%) | 1 (2.3%) | 2 (9.5%) | 0 | 14 (4.0%) | 2 (5.9%) | 0 |
| Patients with HIV/AIDS | 1 (2.6%) | 1 (2.3%) | 2 (9.5%) | 0 | 50 (14.4%) | 5 (14.7%) | 1 (9.1%) |
| Solid organ transplant candidates and recipients | 0 | 1 (2.3%) | 1 (4.8%) | 0 | 9 (2.6%) | 1 (2.9%) | 0 |
| Patients with hematopoietic stem cell transplant | 0 | 1 (2.3%) | 1 (4.8%) | 0 | 8 (2.3%) | 1 (2.9%) | 0 |
| Patients with diabetes mellitus | 4 (10.5%) | 3 (6.8%) | 0 | 0 | 7 (2.0%) | 0 | 0 |
| None of the above | 11 (28.9%) | 16 (36.4%) | 12 (57.1%) | 0 | 126 (36.2%) | 13 (38.2%) | 4 (36.4%) |
| **Total score** | | | | | | | |
| N | 38 | 44 | 21 | 3 | 348 | 34 | 11 |
| Mean | 0.07 | 0.07 | 0.11 | 0.17 | 0.11 | 0.12 | 0.09 |
| Standard deviation | 0.15 | 0.12 | 0.23 | 0.14 | 0.15 | 0.18 | 0.13 |
| Median | 0.00 | 0.00 | 0.00 | 0.25 | 0.00 | 0.00 | 0.00 |
| Minimum | 0.00 | 0.00 | 0.00 | 0.00 | 0.00 | 0.00 | 0.00 |
| Maximum | 0.75 | 0.25 | 1.00 | 0.25 | 0.75 | 0.50 | 0.25 |
| **Quadrivalent HPV, n (%)** | | | | | | | |
| All patients (regardless of age and risk group) | 8 (21.6%) | 8 (22.9%) | 2 (11.8%) | 1 (20.0%) | 86 (17.5%) | 12 (26.7%) | 4 (50.0%) |
| Adults - 20 to 59 years old | 15 (40.5%) | 15 (42.9%) | 2 (11.8%) | 2 (40.0%) | 172 (35.0%) | 17 (37.8%) | 1 (12.5%) |
| Older adults - ≥ 60 years old | 2 (5.4%) | 3 (8.6%) | 0 | 0 | 34 (6.9%) | 1 (2.2%) | 0 |
| Patients with chronic liver disease | 0 | 0 | 0 | 0 | 3 (0.6%) | 0 | 0 |
| Patients with chronic cardiac disease and/or chronic lung disease | 1 (2.7%) | 0 | 0 | 0 | 4 (0.8%) | 0 | 0 |
| Patients with neoplasms or using immunosuppressive drugs | 1 (2.7%) | 1 (2.9%) | 0 | 0 | 13 (2.6%) | 1 (2.2%) | 0 |
| Patients with chronic kidney disease | 0 | 0 | 0 | 0 | 5 (1.0%) | 0 | 0 |

| | | | | | | | |
|---|---|---|---|---|---|---|---|
| Patients with anatomical and functional asplenia | 0 | 0 | 1 (5.9%) | 0 | 3 (0.6%) | 0 | 0 |
| Patients with rheumatic disease | 3 (8.1%) | 0 | 1 (5.9%) | 0 | 5 (1.0%) | 0 | 0 |
| Patients with primary immunodeficiencies | 1 (2.7%) | 1 (2.9%) | 1 (5.9%) | 0 | 20 (4.1%) | 1 (2.2%) | 0 |
| Patients with HIV/AIDS | 2 (5.4%) | 2 (5.7%) | 1 (5.9%) | 0 | 72 (14.7%) | 16 (35.6%) | 0 |
| Solid organ transplant candidates and recipients | 2 (5.4%) | 0 | 1 (5.9%) | 0 | 20 (4.1%) | 1 (2.2%) | 0 |
| Patients with hematopoietic stem cell transplant | 1 (2.7%) | 0 | 1 (5.9%) | 0 | 16 (3.3%) | 1 (2.2%) | 0 |
| Patients with diabetes mellitus | 4 (10.8%) | 2 (5.7%) | 0 | 0 | 8 (1.6%) | 0 | 0 |
| None of the above | 12 (32.4%) | 10 (28.6%) | 12 (70.6%) | 2 (40.0%) | 200 (40.7%) | 11 (24.4%) | 3 (37.5%) |
| **Total score** | | | | | | | |
| N | 37 | 35 | 17 | 5 | 491 | 45 | 8 |
| Mean | 0.09 | 0.10 | 0.03 | 0.10 | 0.10 | 0.17 | 0.03 |
| Standard deviation | 0.12 | 0.14 | 0.08 | 0.14 | 0.15 | 0.21 | 0.09 |
| Median | 0.00 | 0.00 | 0.00 | 0.00 | 0.00 | 0.00 | 0.00 |
| Minimum | 0.00 | 0.00 | 0.00 | 0.00 | 0.00 | 0.00 | 0.00 |
| Maximum | 0.25 | 0.50 | 0.25 | 0.25 | 0.75 | 0.50 | 0.25 |

Note: Proportions were determined based on the number of physicians of each medical specialty who stated to prescribe each vaccine to adult and/or older adult patients.

**Supplementary Table 11 –** Knowledge of vaccines – Dosage schedule in accordance with the recommendations of the Brazilian Society of Immunization by medical specialties

| | Cardiology (n=244) | General Practice/Family Practice (n=85) | Endocrinology (n=80) | Geriatrics (n=26) | Gynecology (n=515) | Infectious Diseases (n=59) | Pulmonology (n=59) |
|---|---|---|---|---|---|---|---|
| **MMR, n (%)** | | | | | | | |
| Single dose | 17 (19.1%) | 15 (25.4%) | 11 (36.7%) | 5 (62.5%) | 90 (28.9%) | 17 (30.9%) | 6 (22.2%) |
| Single dose – the need for additional doses depends on the epidemiological situation | 23 (25.8%) | 17 (28.8%) | 7 (23.3%) | 1 (12.5%) | 54 (17.4%) | 8 (14.5%) | 8 (29.6%) |
| Single annual dose | 1 (1.1%) | 0 | 0 | 0 | 4 (1.3%) | 0 | 0 |
| Two doses | 18 (20.2%) | 15 (25.4%) | 9 (30.0%) | 2 (25.0%) | 92 (29.6%) | 25 (45.5%) | 4 (14.8%) |
| Three doses | 6 (6.7%) | 5 (8.5%) | 1 (3.3%) | 0 | 31 (10.0%) | 1 (1.8%) | 5 (18.5%) |
| Booster dose every 10 years | 13 (14.6%) | 3 (5.1%) | 1 (3.3%) | 0 | 18 (5.8%) | 2 (3.6%) | 2 (7.4%) |
| Sequential schedule with two vaccines (total of three doses) | 3 (3.4%) | 4 (6.8%) | 1 (3.3%) | 0 | 11 (3.5%) | 1 (1.8%) | 1 (3.7%) |
| No need for additional doses | 4 (4.5%) | 0 | 0 | 0 | 1 (0.3%) | 0 | 1 (3.7%) |
| None of the above | 4 (4.5%) | 0 | 0 | 0 | 10 (3.2%) | 1 (1.8%) | 0 |
| Total | 89 | 59 | 30 | 8 | 311 | 55 | 27 |
| **Hepatitis A, n (%)** | | | | | | | |
| Single dose | 27 (32.5%) | 13 (27.7%) | 10 (32.3%) | 2 (18.2%) | 49 (32.5%) | 8 (14.5%) | 7 (38.9%) |
| Single dose – the need for additional doses depends on the epidemiological situation | 13 (15.7%) | 7 (14.9%) | 4 (12.9%) | 2 (18.2%) | 20 (13.2%) | 3 (5.5%) | 3 (16.7%) |
| Single annual dose | 0 | 2 (4.3%) | 1 (3.2%) | 0 | 1 (0.7%) | 0 | 0 |
| Two doses | 20 (24.1%) | 17 (36.2%) | 12 (38.7%) | 4 (36.4%) | 37 (24.5%) | 43 (78.2%) | 5 (27.8%) |
| Three doses | 12 (14.5%) | 3 (6.4%) | 0 | 3 (27.3%) | 25 (16.6%) | 0 | 1 (5.6%) |
| Booster dose every 10 years | 1 (1.2%) | 3 (6.4%) | 1 (3.2%) | 0 | 2 (1.3%) | 0 | 0 |
| Sequential schedule with two vaccines (total of three doses) | 6 (7.2%) | 0 | 3 (9.7%) | 0 | 7 (4.6%) | 0 | 0 |
| No need for additional doses | 1 (1.2%) | 1 (2.1%) | 0 | 0 | 2 (1.3%) | 0 | 1 (5.6%) |
| None of the above | 3 (3.6%) | 1 (2.1%) | 0 | 0 | 8 (5.3%) | 1 (1.8%) | 1 (5.6%) |
| Total | 83 | 47 | 31 | 11 | 151 | 55 | 18 |
| **Hepatitis B, n (%)** | | | | | | | |
| Single dose | 13 (7.0%) | 5 (6.7%) | 3 (4.6%) | 0 | 31 (6.3%) | 1 (1.7%) | 2 (5.4%) |
| Single dose – the need for additional doses depends on the epidemiological situation | 10 (5.3%) | 3 (4.0%) | 0 | 1 (4.3%) | 9 (1.8%) | 0 | 3 (8.1%) |
| Single annual dose | 1 (0.5%) | 1 (1.3%) | 0 | 0 | 1 (0.2%) | 0 | 0 |
| Two doses | 13 (7.0%) | 2 (2.7%) | 2 (3.1%) | 1 (4.3%) | 11 (2.2%) | 1 (1.7%) | 3 (8.1%) |
| Three doses | 122 (65.2%) | 55 (73.3%) | 50 (76.9%) | 18 (78.3%) | 395 (80.8%) | 54 (91.5%) | 24 (64.9%) |
| Booster dose every 10 years | 8 (4.3%) | 3 (4.0%) | 3 (4.6%) | 0 | 6 (1.2%) | 0 | 0 |

| | Cardiology (n=244) | General Practice/Family Practice (n=85) | Endocrinology (n=80) | Geriatrics (n=26) | Gynecology (n=515) | Infectious Diseases (n=59) | Pulmonology (n=59) |
|---|---|---|---|---|---|---|---|
| Sequential schedule with two vaccines (total of three doses) | 17 (9.1%) | 5 (6.7%) | 7 (10.8%) | 3 (13.0%) | 30 (6.1%) | 1 (1.7%) | 4 (10.8%) |
| No need for additional doses | 1 (0.5%) | 1 (1.3%) | 0 | 0 | 3 (0.6%) | 0 | 1 (2.7%) |
| None of the above | 2 (1.1%) | 0 | 0 | 0 | 3 (0.6%) | 2 (3.4%) | 0 |
| Total | 187 | 75 | 65 | 23 | 489 | 59 | 37 |
| **Hepatitis A and B, n (%)** | | | | | | | |
| Single dose | 9 (8.8%) | 9 (16.7%) | 2 (4.9%) | 1 (8.3%) | 18 (7.4%) | 5 (12.2%) | 2 (9.1%) |
| Single dose – the need for additional doses depends on the epidemiological situation | 16 (15.7%) | 3 (5.6%) | 1 (2.4%) | 0 | 12 (4.9%) | 0 | 3 (13.6%) |
| Single annual dose | 2 (2.0%) | 0 | 0 | 0 | 1 (0.4%) | 0 | 0 |
| Two doses | 9 (8.8%) | 6 (11.1%) | 5 (12.2%) | 0 | 20 (8.2%) | 5 (12.2%) | 1 (4.5%) |
| Three doses | 39 (38.2%) | 25 (46.3%) | 18 (43.9%) | 7 (58.3%) | 132 (54.1%) | 22 (53.7%) | 12 (54.5%) |
| Booster dose every 10 years | 9 (8.8%) | 3 (5.6%) | 1 (2.4%) | 0 | 3 (1.2%) | 0 | 0 |
| Sequential schedule with two vaccines (total of three doses) | 8 (7.8%) | 2 (3.7%) | 10 (24.4%) | 3 (25.0%) | 33 (13.5%) | 6 (14.6%) | 1 (4.5%) |
| No need for additional doses | 1 (1.0%) | 1 (1.9%) | 0 | 0 | 2 (0.8%) | 0 | 1 (4.5%) |
| None of the above | 9 (8.8%) | 5 (9.3%) | 4 (9.8%) | 1 (8.3%) | 23 (9.4%) | 3 (7.3%) | 2 (9.1%) |
| Total | 102 | 54 | 41 | 12 | 244 | 41 | 22 |
| **Tdap or Tdap-IPV, n (%)** | | | | | | | |
| Single dose | 21 (19.6%) | 13 (21.0%) | 11 (26.2%) | 4 (20.0%) | 129 (29.3%) | 14 (24.1%) | 10 (31.3%) |
| Single dose – the need for additional doses depends on the epidemiological situation | 20 (18.7%) | 7 (11.3%) | 2 (4.8%) | 4 (20.0%) | 56 (12.7%) | 5 (8.6%) | 5 (15.6%) |
| Single annual dose | 0 | 0 | 0 | 0 | 5 (1.1%) | 0 | 0 |
| Two doses | 7 (6.5%) | 2 (3.2%) | 3 (7.1%) | 1 (5.0%) | 8 (1.8%) | 0 | 4 (12.5%) |
| Three doses | 9 (8.4%) | 10 (16.1%) | 0 | 1 (5.0%) | 66 (15.0%) | 5 (8.6%) | 2 (6.3%) |
| Booster dose every 10 years | 41 (38.3%) | 21 (33.9%) | 21 (50.0%) | 7 (35.0%) | 153 (34.8%) | 31 (53.4%) | 9 (28.1%) |
| Sequential schedule with two vaccines (total of three doses) | 1 (0.9%) | 3 (4.8%) | 1 (2.4%) | 0 | 10 (2.3%) | 1 (1.7%) | 0 |
| No need for additional doses | 2 (1.9%) | 1 (1.6%) | 1 (2.4%) | 0 | 3 (0.7%) | 1 (1.7%) | 1 (3.1%) |
| None of the above | 6 (5.6%) | 5 (8.1%) | 3 (7.1%) | 3 (15.0%) | 10 (2.3%) | 1 (1.7%) | 1 (3.1%) |
| Total | 107 | 62 | 42 | 20 | 440 | 58 | 32 |
| **Tetanus and diphtheria, n (%)** | | | | | | | |
| Single dose | 17 (12.0%) | 8 (11.3%) | 8 (15.4%) | 1 (4.8%) | 47 (10.9%) | 2 (3.5%) | 6 (18.2%) |
| Single dose – the need for additional doses depends on the epidemiological situation | 19 (13.4%) | 7 (9.9%) | 3 (5.8%) | 2 (9.5%) | 19 (4.4%) | 2 (3.5%) | 4 (12.1%) |
| Single annual dose | 2 (1.4%) | 1 (1.4%) | 0 | 0 | 2 (0.5%) | 0 | 0 |
| Two doses | 8 (5.6%) | 6 (8.5%) | 0 | 0 | 11 (2.6%) | 1 (1.8%) | 0 |
| Three doses | 7 (4.9%) | 7 (9.9%) | 0 | 2 (9.5%) | 50 (11.6%) | 1 (1.8%) | 3 (9.1%) |
| Booster dose every 10 years | 81 (57.0%) | 40 (56.3%) | 40 (76.9%) | 15 (71.4%) | 281 (65.3%) | 48 (84.2%) | 18 (54.5%) |

| | Cardiology (n=244) | General Practice/Family Practice (n=85) | Endocrinology (n=80) | Geriatrics (n=26) | Gynecology (n=515) | Infectious Diseases (n=59) | Pulmonology (n=59) |
|---|---|---|---|---|---|---|---|
| Sequential schedule with two vaccines (total of three doses) | 1 (0.7%) | 0 | 1 (1.9%) | 0 | 3 (0.7%) | 0 | 1 (3.0%) |
| No need for additional doses | 1 (0.7%) | 0 | 0 | 0 | 3 (0.7%) | 1 (1.8%) | 1 (3.0%) |
| None of the above | 6 (4.2%) | 2 (2.8%) | 0 | 1 (4.8%) | 14 (3.3%) | 2 (3.5%) | 0 |
| Total | 142 | 71 | 52 | 21 | 430 | 57 | 33 |
| **Varicella, n (%)[a)]** | | | | | | | |
| Single dose | 23 (45.1%) | 16 (38.1%) | 10 (66.7%) | 5 (55.6%) | 48 (45.7%) | 16 (36.4%) | 6 (46.2%) |
| Single dose – the need for additional doses depends on the epidemiological situation | 12 (23.5%) | 5 (11.9%) | 0 | 0 | 14 (13.3%) | 2 (4.5%) | 2 (15.4%) |
| Single annual dose | 2 (3.9%) | 2 (4.8%) | 0 | 0 | 0 | 0 | 0 |
| Two doses | 7 (13.7%) | 15 (35.7%) | 5 (33.3%) | 2 (22.2%) | 34 (32.4%) | 24 (54.5%) | 2 (15.4%) |
| Three doses | 0 | 2 (4.8%) | 0 | 0 | 5 (4.8%) | 0 | 1 (7.7%) |
| Booster dose every 10 years | 4 (7.8%) | 0 | 0 | 0 | 1 (1.0%) | 0 | 0 |
| Sequential schedule with two vaccines (total of three doses) | 1 (2.0%) | 0 | 0 | 0 | 1 (1.0%) | 0 | 0 |
| No need for additional doses | 1 (2.0%) | 0 | 0 | 1 (11.1%) | 1 (1.0%) | 1 (2.3%) | 1 (7.7%) |
| None of the above | 1 (2.0%) | 2 (4.8%) | 0 | 1 (11.1%) | 1 (1.0%) | 1 (2.3%) | 1 (7.7%) |
| Total | 51 | 42 | 15 | 9 | 105 | 44 | 13 |
| **Influenza, n (%)** | | | | | | | |
| Single dose | 31 (13.0%) | 8 (9.8%) | 7 (9.1%) | 2 (7.7%) | 84 (17.0%) | 1 (1.7%) | 10 (17.2%) |
| Single dose – the need for additional doses depends on the epidemiological situation | 18 (7.5%) | 0 | 3 (3.9%) | 0 | 35 (7.1%) | 2 (3.4%) | 4 (6.9%) |
| Single annual dose | 186 (77.8%) | 74 (90.2%) | 67 (87.0%) | 24 (92.3%) | 363 (73.6%) | 55 (93.2%) | 44 (75.9%) |
| Two doses | 1 (0.4%) | 0 | 0 | 0 | 3 (0.6%) | 0 | 0 |
| Three doses | 0 | 0 | 0 | 0 | 0 | 0 | 0 |
| Booster dose every 10 years | 1 (0.4%) | 0 | 0 | 0 | 1 (0.2%) | 0 | 0 |
| Sequential schedule with two vaccines (total of three doses) | 0 | 0 | 0 | 0 | 1 (0.2%) | 0 | 0 |
| No need for additional doses | 0 | 0 | 0 | 0 | 1 (0.2%) | 0 | 0 |
| None of the above | 2 (0.8%) | 0 | 0 | 0 | 5 (1.0%) | 1 (1.7%) | 0 |
| Total | 239 | 82 | 77 | 26 | 493 | 59 | 58 |
| **Meningococcal Conjugate ACWY, n (%)** | | | | | | | |
| Single dose | 14 (29.8%) | 16 (39.0%) | 10 (76.9%) | 0 | 37 (48.7%) | 18 (40.0%) | 7 (43.8%) |
| Single dose – the need for additional doses depends on the epidemiological situation | 13 (27.7%) | 7 (17.1%) | 1 (7.7%) | 1 (25.0%) | 14 (18.4%) | 5 (11.1%) | 4 (25.0%) |
| Single annual dose | 7 (14.9%) | 3 (7.3%) | 0 | 0 | 6 (7.9%) | 0 | 0 |
| Two doses | 6 (12.8%) | 5 (12.2%) | 1 (7.7%) | 1 (25.0%) | 10 (13.2%) | 11 (24.4%) | 3 (18.8%) |
| Three doses | 1 (2.1%) | 4 (9.8%) | 0 | 0 | 3 (3.9%) | 6 (13.3%) | 0 |
| Booster dose every 10 years | 2 (4.3%) | 4 (9.8%) | 1 (7.7%) | 0 | 2 (2.6%) | 1 (2.2%) | 1 (6.3%) |

| | Cardiology (n=244) | General Practice/Family Practice (n=85) | Endocrinology (n=80) | Geriatrics (n=26) | Gynecology (n=515) | Infectious Diseases (n=59) | Pulmonology (n=59) |
|---|---|---|---|---|---|---|---|
| Sequential schedule with two vaccines (total of three doses) | 0 | 1 (2.4%) | 0 | 0 | 1 (1.3%) | 2 (4.4%) | 0 |
| No need for additional doses | 0 | 0 | 0 | 0 | 0 | 0 | 1 (6.3%) |
| None of the above | 4 (8.5%) | 1 (2.4%) | 0 | 2 (50.0%) | 3 (3.9%) | 2 (4.4%) | 0 |
| Total | 47 | 41 | 13 | 4 | 76 | 45 | 16 |
| **Meningococcal B, n (%)[a)]** | | | | | | | |
| Single dose | 14 (31.1%) | 13 (33.3%) | 6 (50.0%) | 1 (25.0%) | 15 (27.8%) | 5 (14.3%) | 4 (33.3%) |
| Single dose – the need for additional doses depends on the epidemiological situation | 14 (31.1%) | 6 (15.4%) | 0 | 1 (25.0%) | 13 (24.1%) | 3 (8.6%) | 3 (25.0%) |
| Single annual dose | 5 (11.1%) | 1 (2.6%) | 0 | 0 | 5 (9.3%) | 0 | 0 |
| Two doses | 5 (11.1%) | 10 (25.6%) | 4 (33.3%) | 0 | 14 (25.9%) | 18 (51.4%) | 3 (25.0%) |
| Three doses | 0 | 3 (7.7%) | 0 | 0 | 3 (5.6%) | 5 (14.3%) | 1 (8.3%) |
| Booster dose every 10 years | 4 (8.9%) | 4 (10.3%) | 1 (8.3%) | 0 | 1 (1.9%) | 2 (5.7%) | 0 |
| Sequential schedule with two vaccines (total of three doses) | 0 | 2 (5.1%) | 0 | 0 | 0 | 1 (2.9%) | 0 |
| No need for additional doses | 0 | 0 | 0 | 1 (25.0%) | 0 | 0 | 1 (8.3%) |
| None of the above | 3 (6.7%) | 0 | 1 (8.3%) | 1 (25.0%) | 3 (5.6%) | 1 (2.9%) | 0 |
| Total | 45 | 39 | 12 | 4 | 54 | 35 | 12 |
| **Yellow Fever, n (%)** | | | | | | | |
| Single dose | 106 (50.2%) | 47 (60.3%) | 43 (66.2%) | 11 (45.8%) | 203 (55.0%) | 49 (83.1%) | 30 (69.8%) |
| Single dose – the need for additional doses depends on the epidemiological situation | 58 (27.5%) | 10 (12.8%) | 10 (15.4%) | 6 (25.0%) | 84 (22.8%) | 6 (10.2%) | 5 (11.6%) |
| Single annual dose | 2 (0.9%) | 1 (1.3%) | 0 | 0 | 2 (0.5%) | 0 | 0 |
| Two doses | 2 (0.9%) | 3 (3.8%) | 0 | 1 (4.2%) | 9 (2.4%) | 3 (5.1%) | 0 |
| Three doses | 1 (0.5%) | 0 | 0 | 0 | 1 (0.3%) | 0 | 0 |
| Booster dose every 10 years | 35 (16.6%) | 16 (20.5%) | 12 (18.5%) | 6 (25.0%) | 62 (16.8%) | 0 | 7 (16.3%) |
| Sequential schedule with two vaccines (total of three doses) | 1 (0.5%) | 0 | 0 | 0 | 0 | 0 | 0 |
| No need for additional doses | 4 (1.9%) | 1 (1.3%) | 0 | 0 | 4 (1.1%) | 1 (1.7%) | 0 |
| None of the above | 2 (0.9%) | 0 | 0 | 0 | 4 (1.1%) | 0 | 1 (2.3%) |
| Total | 211 | 78 | 65 | 24 | 369 | 59 | 43 |
| **PCV13, n (%)[b)]** | | | | | | | |
| Single dose | 95 (48.0%) | 31 (48.4%) | 24 (61.5%) | 17 (68.0%) | 68 (59.1%) | 29 (70.7%) | 42 (77.8%) |
| Single dose – the need for additional doses depends on the epidemiological situation | 21 (10.6%) | 6 (9.4%) | 2 (5.1%) | 3 (12.0%) | 10 (8.7%) | 2 (4.9%) | 4 (7.4%) |
| Single annual dose | 44 (22.2%) | 17 (26.6%) | 5 (12.8%) | 1 (4.0%) | 13 (11.3%) | 1 (2.4%) | 0 |
| Two doses | 14 (7.1%) | 2 (3.1%) | 2 (5.1%) | 1 (4.0%) | 8 (7.0%) | 4 (9.8%) | 2 (3.7%) |
| Three doses | 2 (1.0%) | 1 (1.6%) | 1 (2.6%) | 0 | 4 (3.5%) | 0 | 1 (1.9%) |
| Booster dose every 10 years | 12 (6.1%) | 2 (3.1%) | 2 (5.1%) | 2 (8.0%) | 2 (1.7%) | 2 (4.9%) | 3 (5.6%) |

| | Cardiology (n=244) | General Practice/Family Practice (n=85) | Endocrinology (n=80) | Geriatrics (n=26) | Gynecology (n=515) | Infectious Diseases (n=59) | Pulmonology (n=59) |
|---|---|---|---|---|---|---|---|
| Sequential schedule with two vaccines (total of three doses) | 1 (0.5%) | 2 (3.1%) | 1 (2.6%) | 0 | 4 (3.5%) | 1 (2.4%) | 1 (1.9%) |
| No need for additional doses | 0 | 0 | 0 | 0 | 0 | 0 | 1 (1.9%) |
| None of the above | 9 (4.5%) | 3 (4.7%) | 2 (5.1%) | 1 (4.0%) | 6 (5.2%) | 2 (4.9%) | 0 |
| Total | 198 | 64 | 39 | 25 | 115 | 41 | 54 |
| **PPV23, n (%)[b)]** | | | | | | | |
| Single dose | 83 (43.9%) | 25 (39.7%) | 15 (44.1%) | 8 (30.8%) | 69 (59.0%) | 20 (35.1%) | 22 (37.9%) |
| Single dose – the need for additional doses depends on the epidemiological situation | 20 (10.6%) | 4 (6.3%) | 3 (8.8%) | 4 (15.4%) | 13 (11.1%) | 4 (7.0%) | 5 (8.6%) |
| Single annual dose | 43 (22.8%) | 15 (23.8%) | 3 (8.8%) | 1 (3.8%) | 11 (9.4%) | 0 | 0 |
| Two doses | 13 (6.9%) | 8 (12.7%) | 3 (8.8%) | 9 (34.6%) | 12 (10.3%) | 23 (40.4%) | 10 (17.2%) |
| Three doses | 1 (0.5%) | 1 (1.6%) | 1 (2.9%) | 0 | 4 (3.4%) | 0 | 1 (1.7%) |
| Booster dose every 10 years | 13 (6.9%) | 4 (6.3%) | 3 (8.8%) | 1 (3.8%) | 1 (0.9%) | 3 (5.3%) | 4 (6.9%) |
| Sequential schedule with two vaccines (total of three doses) | 6 (3.2%) | 2 (3.2%) | 2 (5.9%) | 1 (3.8%) | 2 (1.7%) | 2 (3.5%) | 3 (5.2%) |
| No need for additional doses | 3 (1.6%) | 0 | 0 | 0 | 0 | 0 | 0 |
| None of the above | 7 (3.7%) | 4 (6.3%) | 4 (11.8%) | 2 (7.7%) | 5 (4.3%) | 5 (8.8%) | 13 (22.4%) |
| Total | 189 | 63 | 34 | 26 | 117 | 57 | 58 |
| **Zoster, n (%)** | | | | | | | |
| Single dose | 63 (70.8%) | 36 (70.6%) | 27 (87.1%) | 20 (87.0%) | 260 (84.4%) | 32 (78.0%) | 13 (68.4%) |
| Single dose – the need for additional doses depends on the epidemiological situation | 6 (6.7%) | 5 (9.8%) | 0 | 0 | 5 (1.6%) | 0 | 3 (15.8%) |
| Single annual dose | 4 (4.5%) | 0 | 0 | 1 (4.3%) | 2 (0.6%) | 0 | 0 |
| Two doses | 6 (6.7%) | 4 (7.8%) | 3 (9.7%) | 2 (8.7%) | 15 (4.9%) | 8 (19.5%) | 0 |
| Three doses | 1 (1.1%) | 3 (5.9%) | 0 | 0 | 12 (3.9%) | 0 | 1 (5.3%) |
| Booster dose every 10 years | 3 (3.4%) | 2 (3.9%) | 0 | 0 | 3 (1.0%) | 1 (2.4%) | 0 |
| Sequential schedule with two vaccines (total of three doses) | 2 (2.2%) | 0 | 0 | 0 | 1 (0.3%) | 0 | 0 |
| No need for additional doses | 2 (2.2%) | 0 | 0 | 0 | 1 (0.3%) | 0 | 0 |
| None of the above | 2 (2.2%) | 1 (2.0%) | 1 (3.2%) | 0 | 9 (2.9%) | 0 | 2 (10.5%) |
| Total | 89 | 51 | 31 | 23 | 308 | 41 | 19 |
| **Dengue, n (%)[a)]** | | | | | | | |
| Single dose | 18 (32.7%) | 10 (33.3%) | 9 (75.0%) | 1 (33.3%) | 37 (39.8%) | 2 (16.7%) | 2 (25.0%) |
| Single dose – the need for additional doses depends on the epidemiological situation | 22 (40.0%) | 11 (36.7%) | 0 | 1 (33.3%) | 22 (23.7%) | 0 | 1 (12.5%) |
| Single annual dose | 3 (5.5%) | 1 (3.3%) | 0 | 0 | 5 (5.4%) | 1 (8.3%) | 1 (12.5%) |
| Two doses | 1 (1.8%) | 1 (3.3%) | 0 | 0 | 4 (4.3%) | 1 (8.3%) | 0 |
| Three doses | 7 (12.7%) | 4 (13.3%) | 2 (16.7%) | 1 (33.3%) | 15 (16.1%) | 7 (58.3%) | 2 (25.0%) |
| Booster dose every 10 years | 2 (3.6%) | 0 | 0 | 0 | 0 | 0 | 0 |

| | Cardiology (n=244) | General Practice/Family Practice (n=85) | Endocrinology (n=80) | Geriatrics (n=26) | Gynecology (n=515) | Infectious Diseases (n=59) | Pulmonology (n=59) |
|---|---|---|---|---|---|---|---|
| Sequential schedule with two vaccines (total of three doses) | 1 (1.8%) | 2 (6.7%) | 0 | 0 | 1 (1.1%) | 0 | 0 |
| No need for additional doses | 0 | 0 | 0 | 0 | 1 (1.1%) | 0 | 0 |
| None of the above | 1 (1.8%) | 1 (3.3%) | 1 (8.3%) | 0 | 8 (8.6%) | 1 (8.3%) | 2 (25.0%) |
| Total | 55 | 30 | 12 | 3 | 93 | 12 | 8 |
| **Bivalent HPV, n (%)[a]** | | | | | | | |
| Single dose | 18 (47.4%) | 13 (29.5%) | 4 (19.0%) | 1 (33.3%) | 19 (5.5%) | 2 (5.9%) | 1 (9.1%) |
| Single dose – the need for additional doses depends on the epidemiological situation | 4 (10.5%) | 3 (6.8%) | 0 | 0 | 3 (0.9%) | 0 | 0 |
| Single annual dose | 0 | 0 | 0 | 0 | 1 (0.3%) | 0 | 0 |
| Two doses | 6 (15.8%) | 10 (22.7%) | 8 (38.1%) | 0 | 65 (18.7%) | 10 (29.4%) | 4 (36.4%) |
| Three doses | 5 (13.2%) | 13 (29.5%) | 7 (33.3%) | 2 (66.7%) | 228 (65.5%) | 19 (55.9%) | 4 (36.4%) |
| Booster dose every 10 years | 0 | 1 (2.3%) | 0 | 0 | 0 | 0 | 0 |
| Sequential schedule with two vaccines (total of three doses) | 1 (2.6%) | 1 (2.3%) | 1 (4.8%) | 0 | 20 (5.7%) | 0 | 1 (9.1%) |
| No need for additional doses | 1 (2.6%) | 0 | 0 | 0 | 1 (0.3%) | 0 | 0 |
| None of the above | 3 (7.9%) | 3 (6.8%) | 1 (4.8%) | 0 | 11 (3.2%) | 3 (8.8%) | 1 (9.1%) |
| Total | 38 | 44 | 21 | 3 | 348 | 34 | 11 |
| **Quadrivalent HPV, n (%)[a]** | | | | | | | |
| Single dose | 13 (35.1%) | 14 (40.0%) | 4 (23.5%) | 0 | 18 (3.7%) | 2 (4.4%) | 2 (25.0%) |
| Single dose – the need for additional doses depends on the epidemiological situation | 4 (10.8%) | 2 (5.7%) | 0 | 0 | 4 (0.8%) | 0 | 1 (12.5%) |
| Single annual dose | 0 | 0 | 0 | 0 | 1 (0.2%) | 0 | 0 |
| Two doses | 6 (16.2%) | 10 (28.6%) | 2 (11.8%) | 0 | 94 (19.1%) | 9 (20.0%) | 2 (25.0%) |
| Three doses | 8 (21.6%) | 8 (22.9%) | 9 (52.9%) | 4 (80.0%) | 330 (67.2%) | 31 (68.9%) | 2 (25.0%) |
| Booster dose every 10 years | 1 (2.7%) | 1 (2.9%) | 0 | 0 | 1 (0.2%) | 0 | 0 |
| Sequential schedule with two vaccines (total of three doses) | 2 (5.4%) | 0 | 1 (5.9%) | 0 | 27 (5.5%) | 0 | 1 (12.5%) |
| No need for additional doses | 0 | 0 | 0 | 0 | 1 (0.2%) | 0 | 0 |
| None of the above | 3 (8.1%) | 0 | 1 (5.9%) | 1 (20.0%) | 15 (3.1%) | 3 (6.7%) | 0 |
| Total | 37 | 35 | 17 | 5 | 491 | 45 | 8 |

[a] For physicians who prescribed the vaccine to adult patients.

[b] For physicians who prescribed the vaccine to older adult patients.

MMR, measle, mumps, and rubella; Tdap, adult tetanus-diphtheria-acellular; PCV13, 13-valent pneumococcal conjugate vaccine; PPV23, 23-valent pneumococcal polysaccharide vaccine; HPV, human papillomavirus.

Note: Proportions were determined based on the number of physicians of each medical specialty who stated to prescribe each vaccine to adult and/or older adult patients.

Correct answers are highlighted.

**Supplementary Table 12** – Knowledge of vaccines – Availability in Brazil's Unified Public Health System (SUS) – free of charge at the Basic Health Units by medical specialties

| | Cardiology (n=244) | General Practice/Family Practice (n=85) | Endocrinology (n=80) | Geriatrics (n=26) | Gynecology (n=515) | Infectious Diseases (n=59) | Pulmonology (n=59) |
|---|---|---|---|---|---|---|---|
| **MMR (measles, mumps and rubella), n (%)** | | | | | | | |
| Yes, in adults and older adults | 52 (58.4%) | 29 (49.2%) | 16 (53.3%) | 6 (75.0%) | 151 (48.6%) | 19 (34.5%) | 17 (63.0%) |
| Yes, only in adults | 19 (21.3%) | 23 (39.0%) | 13 (43.3%) | 2 (25.0%) | 116 (37.3%) | 33 (60.0%) | 10 (37.0%) |
| Yes, only in older adults | 2 (2.2%) | 0 | 0 | 0 | 2 (0.6%) | 0 | 0 |
| Nor in adults or older adults | 16 (18.0%) | 7 (11.9%) | 1 (3.3%) | 0 | 42 (13.5%) | 3 (5.5%) | 0 |
| Total | 89 | 59 | 30 | 8 | 311 | 55 | 27 |
| **Hepatitis A, n (%)** | | | | | | | |
| Yes, in adults and older adults | 47 (56.6%) | 26 (55.3%) | 15 (48.4%) | 4 (36.4%) | 50 (33.1%) | 18 (32.7%) | 8 (44.4%) |
| Yes, only in adults | 10 (12.0%) | 7 (14.9%) | 2 (6.5%) | 4 (36.4%) | 20 (13.2%) | 10 (18.2%) | 5 (27.8%) |
| Yes, only in older adults | 1 (1.2%) | 2 (4.3%) | 0 | 0 | 1 (0.7%) | 0 | 0 |
| Nor in adults or older adults | 25 (30.1%) | 12 (25.5%) | 14 (45.2%) | 3 (27.3%) | 80 (53.0%) | 27 (49.1%) | 5 (27.8%) |
| Total | 83 | 47 | 31 | 11 | 151 | 55 | 18 |
| **Hepatitis B, n (%)** | | | | | | | |
| Yes, in adults and older adults | 145 (77.5%) | 57 (76.0%) | 47 (72.3%) | 19 (82.6%) | 338 (69.1%) | 45 (76.3%) | 28 (75.7%) |
| Yes, only in adults | 23 (12.3%) | 13 (17.3%) | 9 (13.8%) | 4 (17.4%) | 115 (23.5%) | 12 (20.3%) | 6 (16.2%) |
| Yes, only in older adults | 4 (2.1%) | 1 (1.3%) | 1 (1.5%) | 0 | 4 (0.8%) | 0 | 1 (2.7%) |
| Nor in adults or older adults | 15 (8.0%) | 4 (5.3%) | 8 (12.3%) | 0 | 32 (6.5%) | 2 (3.4%) | 2 (5.4%) |
| Total | 187 | 75 | 65 | 23 | 489 | 59 | 37 |
| **Hepatitis A and B, n (%)** | | | | | | | |
| Yes, in adults and older adults | 53 (52.0%) | 18 (33.3%) | 11 (26.8%) | 8 (66.7%) | 83 (34.0%) | 9 (22.0%) | 11 (50.0%) |
| Yes, only in adults | 9 (8.8%) | 8 (14.8%) | 4 (9.8%) | 0 | 39 (16.0%) | 3 (7.3%) | 5 (22.7%) |
| Yes, only in older adults | 5 (4.9%) | 2 (3.7%) | 1 (2.4%) | 0 | 6 (2.5%) | 0 | 1 (4.5%) |
| Nor in adults or older adults | 35 (34.3%) | 26 (48.1%) | 25 (61.0%) | 4 (33.3%) | 116 (47.5%) | 29 (70.7%) | 5 (22.7%) |
| Total | 102 | 54 | 41 | 12 | 244 | 41 | 22 |
| **Adult triple acellular bacterial vaccine (diphtheria, tetanus and acellular pertussis) - Tdap or Tdap-IPV, n (%)** | | | | | | | |
| Yes, in adults and older adults | 71 (66.4%) | 31 (50.0%) | 24 (57.1%) | 11 (55.0%) | 256 (58.2%) | 19 (32.8%) | 18 (56.3%) |
| Yes, only in adults | 15 (14.0%) | 12 (19.4%) | 4 (9.5%) | 4 (20.0%) | 109 (24.8%) | 12 (20.7%) | 5 (15.6%) |
| Yes, only in older adults | 3 (2.8%) | 3 (4.8%) | 4 (9.5%) | 0 | 4 (0.9%) | 0 | 3 (9.4%) |
| Nor in adults or older adults | 18 (16.8%) | 16 (25.8%) | 10 (23.8%) | 5 (25.0%) | 71 (16.1%) | 27 (46.6%) | 6 (18.8%) |
| Total | 107 | 62 | 42 | 20 | 440 | 58 | 32 |
| **Tetanus and diphtheria (diphtheria and tetanus) - dT, n (%)** | | | | | | | |
| Yes, in adults and older adults | 118 (83.1%) | 55 (77.5%) | 48 (92.3%) | 20 (95.2%) | 342 (79.5%) | 54 (94.7%) | 26 (78.8%) |

| | Cardiology (n=244) | General Practice/Family Practice (n=85) | Endocrinology (n=80) | Geriatrics (n=26) | Gynecology (n=515) | Infectious Diseases (n=59) | Pulmonology (n=59) |
|---|---|---|---|---|---|---|---|
| Yes, only in adults | 13 (9.2%) | 9 (12.7%) | 2 (3.8%) | 1 (4.8%) | 69 (16.0%) | 2 (3.5%) | 5 (15.2%) |
| Yes, only in older adults | 5 (3.5%) | 1 (1.4%) | 0 | 0 | 2 (0.5%) | 0 | 1 (3.0%) |
| Nor in adults or older adults | 6 (4.2%) | 6 (8.5%) | 2 (3.8%) | 0 | 17 (4.0%) | 1 (1.8%) | 1 (3.0%) |
| Total | 142 | 71 | 52 | 21 | 430 | 57 | 33 |
| **Varicella (Chickenpox), n (%)** | | | | | | | |
| Yes, in adults and older adults | 24 (47.1%) | 14 (33.3%) | 6 (40.0%) | 2 (22.2%) | 28 (26.7%) | 12 (27.3%) | 5 (38.5%) |
| Yes, only in adults | 10 (19.6%) | 4 (9.5%) | 1 (6.7%) | 1 (11.1%) | 27 (25.7%) | 8 (18.2%) | 3 (23.1%) |
| Yes, only in older adults | 1 (2.0%) | 1 (2.4%) | 1 (6.7%) | 0 | 3 (2.9%) | 1 (2.3%) | 0 |
| Nor in adults or older adults | 16 (31.4%) | 23 (54.8%) | 7 (46.7%) | 6 (66.7%) | 47 (44.8%) | 23 (52.3%) | 5 (38.5%) |
| Total | 51 | 42 | 15 | 9 | 105 | 44 | 13 |
| **Influenza (flu), n (%)** | | | | | | | |
| Yes, in adults and older adults | 151 (63.2%) | 53 (64.6%) | 47 (61.0%) | 20 (76.9%) | 365 (74.0%) | 41 (69.5%) | 30 (51.7%) |
| Yes, only in adults | 4 (1.7%) | 5 (6.1%) | 1 (1.3%) | 0 | 8 (1.6%) | 1 (1.7%) | 0 |
| Yes, only in older adults | 83 (34.7%) | 22 (26.8%) | 29 (37.7%) | 6 (23.1%) | 116 (23.5%) | 16 (27.1%) | 28 (48.3%) |
| Nor in adults or older adults | 1 (0.4%) | 2 (2.4%) | 0 | 0 | 4 (0.8%) | 1 (1.7%) | 0 |
| Total | 239 | 82 | 77 | 26 | 493 | 59 | 58 |
| **Meningococcal Conjugate ACWY, n (%)** | | | | | | | |
| Yes, in adults and older adults | 24 (51.1%) | 13 (31.7%) | 3 (23.1%) | 1 (25.0%) | 24 (31.6%) | 10 (22.2%) | 7 (43.8%) |
| Yes, only in adults | 4 (8.5%) | 6 (14.6%) | 0 | 1 (25.0%) | 19 (25.0%) | 3 (6.7%) | 4 (25.0%) |
| Yes, only in older adults | 3 (6.4%) | 1 (2.4%) | 0 | 0 | 1 (1.3%) | 0 | 0 |
| Nor in adults or older adults | 16 (34.0%) | 21 (51.2%) | 10 (76.9%) | 2 (50.0%) | 32 (42.1%) | 32 (71.1%) | 5 (31.3%) |
| Total | 47 | 41 | 13 | 4 | 76 | 45 | 16 |
| **Meningococcal B, n (%)** | | | | | | | |
| Yes, in adults and older adults | 27 (60.0%) | 15 (38.5%) | 2 (16.7%) | 2 (50.0%) | 22 (40.7%) | 5 (14.3%) | 5 (41.7%) |
| Yes, only in adults | 6 (13.3%) | 3 (7.7%) | 0 | 1 (25.0%) | 10 (18.5%) | 3 (8.6%) | 2 (16.7%) |
| Yes, only in older adults | 2 (4.4%) | 1 (2.6%) | 0 | 0 | 2 (3.7%) | 0 | 0 |
| Nor in adults or older adults | 10 (22.2%) | 20 (51.3%) | 10 (83.3%) | 1 (25.0%) | 20 (37.0%) | 27 (77.1%) | 5 (41.7%) |
| Total | 45 | 39 | 12 | 4 | 54 | 35 | 12 |
| **Yellow Fever, n (%)** | | | | | | | |
| Yes, in adults and older adults | 175 (82.9%) | 62 (79.5%) | 53 (81.5%) | 19 (79.2%) | 301 (81.6%) | 35 (59.3%) | 36 (83.7%) |
| Yes, only in adults | 32 (15.2%) | 15 (19.2%) | 12 (18.5%) | 4 (16.7%) | 65 (17.6%) | 22 (37.3%) | 6 (14.0%) |
| Yes, only in older adults | 2 (0.9%) | 0 | 0 | 0 | 1 (0.3%) | 0 | 0 |
| Nor in adults or older adults | 2 (0.9%) | 1 (1.3%) | 0 | 1 (4.2%) | 2 (0.5%) | 2 (3.4%) | 1 (2.3%) |
| Total | 211 | 78 | 65 | 24 | 369 | 59 | 43 |
| **Anti-pneumococcal conjugated 13-valent vaccine, n (%)** | | | | | | | |
| Yes, in adults and older adults | 71 (34.1%) | 27 (39.7%) | 16 (36.4%) | 9 (36.0%) | 66 (50.4%) | 10 (21.3%) | 21 (36.8%) |

| | Cardiology (n=244) | General Practice/Family Practice (n=85) | Endocrinology (n=80) | Geriatrics (n=26) | Gynecology (n=515) | Infectious Diseases (n=59) | Pulmonology (n=59) |
|---|---|---|---|---|---|---|---|
| Yes, only in adults | 5 (2.4%) | 1 (1.5%) | 0 | 0 | 4 (3.1%) | 1 (2.1%) | 2 (3.5%) |
| Yes, only in older adults | 47 (22.6%) | 14 (20.6%) | 10 (22.7%) | 3 (12.0%) | 25 (19.1%) | 3 (6.4%) | 4 (7.0%) |
| Nor in adults or older adults | 85 (40.9%) | 26 (38.2%) | 18 (40.9%) | 13 (52.0%) | 36 (27.5%) | 33 (70.2%) | 30 (52.6%) |
| Total | 208 | 68 | 44 | 25 | 131 | 47 | 57 |
| **Anti-pneumococcal conjugated 23-valent vaccine, n (%)** | | | | | | | |
| Yes, in adults and older adults | 66 (33.8%) | 21 (31.8%) | 8 (21.6%) | 10 (38.5%) | 66 (52.0%) | 23 (39.7%) | 30 (51.7%) |
| Yes, only in adults | 4 (2.1%) | 3 (4.5%) | 0 | 0 | 4 (3.1%) | 2 (3.4%) | 2 (3.4%) |
| Yes, only in older adults | 42 (21.5%) | 19 (28.8%) | 7 (18.9%) | 9 (34.6%) | 23 (18.1%) | 17 (29.3%) | 17 (29.3%) |
| Nor in adults or older adults | 83 (42.6%) | 23 (34.8%) | 22 (59.5%) | 7 (26.9%) | 34 (26.8%) | 16 (27.6%) | 9 (15.5%) |
| Total | 195 | 66 | 37 | 26 | 127 | 58 | 58 |
| **Zoster, n (%)** | | | | | | | |
| Yes, in adults and older adults | 32 (36.0%) | 13 (25.5%) | 6 (19.4%) | 3 (13.0%) | 84 (27.3%) | 7 (17.1%) | 8 (42.1%) |
| Yes, only in adults | 2 (2.2%) | 5 (9.8%) | 0 | 0 | 14 (4.5%) | 0 | 1 (5.3%) |
| Yes, only in older adults | 8 (9.0%) | 6 (11.8%) | 2 (6.5%) | 2 (8.7%) | 60 (19.5%) | 3 (7.3%) | 0 |
| Nor in adults or older adults | 47 (52.8%) | 27 (52.9%) | 23 (74.2%) | 18 (78.3%) | 150 (48.7%) | 31 (75.6%) | 10 (52.6%) |
| Total | 89 | 51 | 31 | 23 | 308 | 41 | 19 |
| **Dengue, n (%)** | | | | | | | |
| Yes, in adults and older adults | 40 (72.7%) | 19 (63.3%) | 7 (58.3%) | 2 (66.7%) | 44 (47.3%) | 3 (25.0%) | 6 (75.0%) |
| Yes, only in adults | 5 (9.1%) | 1 (3.3%) | 1 (8.3%) | 1 (33.3%) | 12 (12.9%) | 1 (8.3%) | 0 |
| Yes, only in older adults | 0 | 0 | 0 | 0 | 0 | 0 | 0 |
| Nor in adults or older adults | 10 (18.2%) | 10 (33.3%) | 4 (33.3%) | 0 | 37 (39.8%) | 8 (66.7%) | 2 (25.0%) |
| Total | 55 | 30 | 12 | 3 | 93 | 12 | 8 |
| **Bivalent Fever, n (%)** | | | | | | | |
| Yes, in adults and older adults | 14 (36.8%) | 9 (20.5%) | 2 (9.5%) | 0 | 57 (16.4%) | 1 (2.9%) | 2 (18.2%) |
| Yes, only in adults | 8 (21.1%) | 14 (31.8%) | 2 (9.5%) | 2 (66.7%) | 95 (27.3%) | 7 (20.6%) | 3 (27.3%) |
| Yes, only in older adults | 0 | 0 | 1 (4.8%) | 0 | 1 (0.3%) | 1 (2.9%) | 0 |
| Nor in adults or older adults | 16 (42.1%) | 21 (47.7%) | 16 (76.2%) | 1 (33.3%) | 195 (56.0%) | 25 (73.5%) | 6 (54.5%) |
| Total | 38 | 44 | 21 | 3 | 348 | 34 | 11 |
| **Quadrivalent HPV, n (%)** | | | | | | | |
| Yes, in adults and older adults | 10 (27.0%) | 9 (25.7%) | 2 (11.8%) | 1 (20.0%) | 67 (13.6%) | 3 (6.7%) | 2 (25.0%) |
| Yes, only in adults | 9 (24.3%) | 10 (28.6%) | 3 (17.6%) | 1 (20.0%) | 135 (27.5%) | 15 (33.3%) | 4 (50.0%) |
| Yes, only in older adults | 1 (2.7%) | 0 | 0 | 0 | 4 (0.8%) | 0 | 0 |
| Nor in adults or older adults | 17 (45.9%) | 16 (45.7%) | 12 (70.6%) | 3 (60.0%) | 285 (58.0%) | 27 (60.0%) | 2 (25.0%) |
| Total | 37 | 35 | 17 | 5 | 491 | 45 | 8 |

| | Cardiology (n=244) | General Practice/Family Practice (n=85) | Endocrinology (n=80) | Geriatrics (n=26) | Gynecology (n=515) | Infectious Diseases (n=59) | Pulmonology (n=59) |
|---|---|---|---|---|---|---|---|

Note: Proportions were determined based on the number of physicians of each medical specialty who stated to prescribe each vaccine to adult and/or older adult patients. Correct answers are highlighted.

**Supplementary Table 13** – Comparison of the populations of physicians in Brazil, invited to participate in the study, and completing the survey

| | Brazil** | Invited Physicians*** | Survey |
|---|---|---|---|
| Number of physicians, n | 452 801 | 76 104 | 1068 |
| Sex (female), n (%) | 45.6% | 36.7% | 53.7% |
| Age (mean) | 45.4 years | Not available | 44.70 years |
| Region, % | | | |
| North | 4.6% | 2.2% | 4.5% |
| Northeast | 17.8% | 11.3% | 17.9% |
| Midwest | 8.3% | 5.0% | 8.2% |
| Southeast | 54.1% | 70.8% | 54.3% |
| South | 15.1% | 10.6% | 15.1% |
| Primary area of practice, n (%) | | | |
| Public | 21.6% | Not available | 4.8% |
| Private | 26.9% | Not available | 33.6% |
| Public-private | 51.5% | Not available | 61.6% |
| Medical specialty*, n (%) | | | |
| Cardiology | 13420 (22.9%) | 14662 (19.3%) | 244 (22.8%) |
| GP/FP | 4682 (8.0%) | 35697 (46.9%) | 85 (8.0%) |
| Geriatrics | 1405 (2.4%) | 1737 (2.3%) | 26 (2.4%) |
| Gynecology | 28280 (48.2%) | 13259 (17.4%) | 515 (48.2%) |
| Endocrinology | 4396 (7.5%) | 4166 (5.5%) | 80 (7.5%) |
| Pulmonology | 3253 (5.5%) | 3749 (4.9%) | 59 (5.5%) |
| Infectious Diseases | 3229 (5.5%) | 2834 (3.7%) | 59 (5.5%) |

*Percentages (%) correspond to the number of physicians in each medical specialty divided by the sum of physicians in each medical specialty of interest.

**According to Mário Scheffer et al. Demografia Médica no Brasil 2015. [Internet] 2015 [Accessed on 06 Set 2017]. Available at: https://www.usp.br/agen/wpcontent/uploads/DemografiaMedica30nov2015.pdf.

***Includes all invited physicians – those already registered in the Fine Panel at the time of recruitment and those who registered in the panel only after being invited to complete the survey.

**Sample size stratification**

The number of physicians required to complete the survey per region was estimated based on the number of active physicians in Brazil (total and per region) (4), as presented below.

| Region | Active Physicians | % for stratification* | Total number of required answers** |
|---|---|---|---|
| North | 21584 | 4 | 48 |
| Northeast | 85954 | 18 | 191 |
| Midwest | 39508 | 8 | 88 |
| Southeast | 261852 | 54 | 581 |
| South | 72482 | 15 | 161 |
| Total | 481380 | 100 | 1068 |

*Rational: Number of active physicians in the region/ total of active physicians in Brazil (4)

**Rational: % for stratification X number of required answers to the survey (1068)

On the other hand, the number of physicians required to complete the survey per medical specialty was estimated, based on the number of physicians of each medical specialty to be included in the study in Brazil (5). This process was followed for each medical specialty except General Practice. This is because the number of physicians belonging exclusively to General Practice is reduced in Brazil, and it was expected to be considerably lower than the number presented in the reference used (in which physicians with more than one medical specialty were considered) (5).

As such, to estimate the number of physicians working exclusively as general practitioners, a different procedure was followed. Initially, based on the Regional Council of Medicine of São Paulo's website (6), the number of active physicians working exclusively as general practitioners was determined, and the proportion of these physicians in São Paulo was calculated. Subsequently, the number of general practitioners in the remaining Southeast region's states was determined, assuming the same proportion found in São Paulo. Finally, the proportion of general practitioners in the Southeast region was estimated and used to assess the number of physicians belonging exclusively to this specialty in the remaining Brazilian regions, which allowed to obtain the total number in the country.

| Specialty | Physicians per medical specialty | % for stratification* | Total number of required answers** |
|---|---|---|---|
| **Cardiology** | 13420 | 23 | 244 |
| **GP/FP** | 4682 | 8 | 85 |
| **Endocrinology** | 4396 | 7 | 80 |
| **Geriatrics** | 1405 | 2 | 26 |
| **Gynecology** | 28280 | 48 | 515 |
| **ID** | 3229 | 6 | 59 |
| **Pulmonology** | 3253 | 6 | 59 |
| **Total** | 87205 | 100 | 1068 |

GP/FP, general practitioners/family physicians.

*Rational: Number of physicians from each specialty/ total number of physicians from the medical specialties to be included in the study.

**Rational: % for stratification X number of required answers to the survey (1068)

Finally, based on this information, the number of survey answers from physicians of each medical specialty per Brazilian region was determined.
